# Multimodal macroscopic brain abnormalities in schizophrenia converge on insular, mediofrontal, and sensorimotor cortices and link with glial cell expression but not with recent evolutionary change

**DOI:** 10.64898/2026.09.30.26364383

**Authors:** Melissa Thalhammer, Paula Roßmüller, Chun Meng, Josef Priller, Christian Sorg, Felix Brandl

## Abstract

**Background:** Schizophrenia is characterized by widespread structural and functional brain abnormalities, yet it remains unclear whether these alterations converge across imaging modalities and whether such convergence is associated with underlying cellular distribution and evolution of the human cortex.

**Methods:** We performed coordinate-based meta-analyses of seven whole-brain neuroimaging modalities, including cortical thickness, gyrification, voxel-based morphometry, amplitude of low-frequency fluctuations, regional homogeneity, regional cerebral blood flow (all based on magnetic resonance imaging), and glucose metabolism based on [^18^F]-Fluorodeoxyglucose positron emission tomography. The analyses included 200 studies (209 datasets) comprising 11,794 patients with schizophrenia and 10,342 healthy controls. After single-modality meta-analyses using seed-based d-mapping, the meta-analytic result maps were integrated using orthonormal projective non-negative matrix factorization. Finally, the resulting component maps were related to maps of cortical cell type-specific gene expression and human-specific cortical evolution.

**Results:** Multimodal integration identified a robust convergence pattern centered in bilateral insular, mediofrontal, and lateral sensorimotor cortices, with the strongest contributions from regional homogeneity, cerebral blood flow, and amplitude of low-frequency fluctuations. The convergence pattern showed significant spatial correspondence with cortical gene expression profiles of astrocytes, microglia, and oligodendrocyte progenitor cells. In contrast, we found no evidence for a preferential localization to regions of human-specific cortical expansion or connectivity.

**Conclusions:** Structural and functional brain abnormalities in schizophrenia converge on an insular-mediofrontal-sensorimotor pattern that aligns with glial gene expression profiles. These results support glia-related regional vulnerability as an organizing principle of multimodal brain abnormalities in schizophrenia.

## Introduction

Schizophrenia is a debilitating psychiatric disorder characterized by substantial heterogeneity across multiple levels of organization, ranging from risk factors (1) and clinical symptoms to macroscopic brain alterations (2–4) and cellular underpinnings (5–7). Neuroimaging studies have consistently demonstrated widespread structural and functional brain abnormalities in schizophrenia (8–14). However, their spatial distribution and magnitude vary considerably across patients (15,16), brain regions, and imaging modalities (2,3).

Although this multi-scale heterogeneity is increasingly recognized, most neuroimaging studies focus on single imaging modalities and relatively small samples, limiting our understanding of relationships across modalities and biological scales (17–20). To address this limitation, multimodal approaches have begun to integrate findings across imaging techniques to determine whether macroscopic brain alterations converge on shared spatial patterns (2,21,22). In parallel, advances in multiscale neuroscience (23–25) make it possible to investigate whether these convergent imaging abnormalities reflect underlying biological processes, such as neurotransmitter systems or cellular architecture (21,22,26). Recent work integrating four structural and functional imaging modalities identified convergent abnormalities in the insula, anterior cingulate, frontotemporal cortex, and subcortical regions (21), corroborating theories that schizophrenia involves coordinated disruptions across multiple aspects of brain organization rather than isolated modality-specific alterations (17). Furthermore, these regions were associated with the spatial distribution of specific neurotransmitter systems, including dopaminergic and glutamatergic pathways (21). However, it remains unclear whether such convergence extends to additional imaging modalities and whether it reflects broader principles of cortical organization, such as expression of specific cell types (as an extension to neurotransmitter systems (21)) and recent evolutionary change.

Regarding cellular architecture, postmortem and genetic studies suggest that schizophrenia involves diverse cellular and molecular underpinnings, encompassing both neuronal (6,7,27) and glial processes (5,28,29). While common genetic risk is concentrated in neurons (7,30), complementary models propose that impaired glial progenitor maturation might contribute to disease manifestation (5). Consistent with this view, studies using imaging transcriptomics have linked macroscale brain abnormalities to regional gene expression profiles of glial cell types (26,31).

Another longstanding hypothesis proposes that the evolution of higher-order cognitive functions, social cognition, and language has increased the vulnerability for distortion of these complex functions, manifesting in disorders like schizophrenia (32,33). Supporting this view, schizophrenia genetic risk loci are enriched in or near human accelerated regions (HARs), i.e., genomic regions that have remained highly conserved in other species but underwent rapid change in the human lineage (34). Previous neuroimaging studies also showed that altered brain connectivity - a prominent feature of schizophrenia (35–38) - disproportionately affects connections that are specific to humans compared to our closest living relative, the chimpanzee (39).

In the present study, we investigated whether macroscopic brain abnormalities in schizophrenia converge across multiple neuroimaging modalities and whether these convergence patterns are associated with cell-type expression patterns and recent evolutionary change, as exemplary and complementary instances of further biological characterization. We extended previous work (21) by increasing the number of investigated neuroimaging modalities, by employing an advanced multimodal integration approach, and by linking these findings to both cell distribution and human evolution. First, we conducted separate coordinate-based meta-analyses of whole-brain imaging studies that investigated abnormalities in patients with schizophrenia compared to healthy controls regarding cortical thickness (CTh), gyrification, voxel-based morphometry (VBM) with modulation interpreted as gray matter volume (GMV), amplitude of low-frequency fluctuations (ALFF), regional homogeneity (ReHo), arterial spin labelling-derived regional cerebral blood flow (rCBF) – all derived from magnetic resonance imaging –, and glucose metabolism derived from [^18^F]-Fluorodeoxyglucose positron emission tomography (FDG-PET). Next, to test for convergence of multimodal abnormalities, we integrated the meta-analytic result maps using non-negative matrix factorization (NMF), a parts-based dimensionality reduction approach that identifies latent patterns of covariance across modalities (40–43). Finally, we tested whether the resulting convergence patterns were associated with cortical cell-type distributions and markers of human-specific cortical evolution using previously published reference maps (34,39,44).

## Methods and Materials

### Coordinate-based meta-analyses of macroscopic brain imaging abnormalities

#### Systematic literature search and study selection

We followed the Preferred Reporting Items for Systematic Reviews and Meta-Analyses (PRISMA) guidelines (45,46). The study protocol was registered in the International Prospective Register of Systematic Reviews (PROSPERO, registration no. CRD420261288726). PubMed was systematically searched for cross-sectional whole-brain studies published until January 1, 2026, that compared patients with schizophrenia and healthy controls using the seven imaging modalities listed above. These modalities were selected for their uni-regional outcome measures in contrast to, for example, connectivity outcomes, which are at least bi-regional and therefore harder to examine with coordinate-based meta-analyses. Two researchers (MT and PR) independently screened studies, extracted data, and resolved discrepancies by consensus with a third researcher (FB). Detailed search strategies, eligibility criteria, quality assessment, and data extraction procedures are provided in the Supplementary Methods and Supplementary Figure S1.

#### Single-modality meta-analyses

We conducted seven separate meta-analyses, one for each modality, using Seed-based *d* Mapping with Permutation of Subject Images (SDM-PSI) (version 6.23; https://www.sdmproject.com/software/ (47)). SDM-PSI reconstructs study-specific voxel-wise effect-size maps from reported peak coordinates and synthesizes them using random-effects meta-analysis weighted by sample size and between-study heterogeneity. Statistical significance was defined as p<0.001. Furthermore, we evaluated heterogeneity and publication bias (48,49) and calculated the percentage of variability that reflected the residual heterogeneity across studies (I^2^ statistic) (50). For details, see Supplementary Methods.

### Multimodal integration of single-modality meta-analyses results using NMF

To investigate the spatial convergence of structural and functional brain abnormalities in schizophrenia, we applied the orthonormal projective variant of NMF (OPNMF) to the unthresholded single-modality meta-analytic z-maps. OPNMF is a parts-based dimensionality reduction approach that decomposes multimodal data into spatial components and modality-specific weights while constraining solutions to be non-negative, facilitating biological interpretation (40–42). For details, see Supplementary Methods.

The analyses were performed using publicly available MATLAB code (https://github.com/asotiras/brainparts/blob/master/opnmf_mem.m) with default parameters (non-negative double singular value decomposition (NNDSVD) initialization, max iterations = 50,000, tolerance = 1e-5) in MATLAB 2023b (51) on a MacBook Pro with Apple M2 chip and 16 GB of RAM. To control our approach, we used principal component analysis (PCA) as an alternative multimodal integration strategy in supplementary analyses (Supplementary Methods). Since CTh and gyrification are restricted to cortical voxels (i.e., SDM’s “Freesurfer mask”), the resulting component maps were inherently restricted to cortical voxels.

To statistically evaluate voxel-wise contributions to the OPNMF-derived component maps, we used Brain Surrogate Maps with Autocorrelated Spatial Heterogeneity (BrainSMASH; https://brainsmash.readthedocs.io/en/latest/index.html), which generates surrogate brain maps that preserve the spatial autocorrelation of the original data while randomizing the spatial distribution of values (52). To generate voxel-wise null distributions for subsequent empirical p-value calculation, OPNMF was applied to the original data and to 1,000 surrogate maps per modality, generated with BrainSMASH. Voxels with p < 0.05 were retained in the final component maps. For details, see Supplementary Methods and Fig. 1.

**Figure 1:**
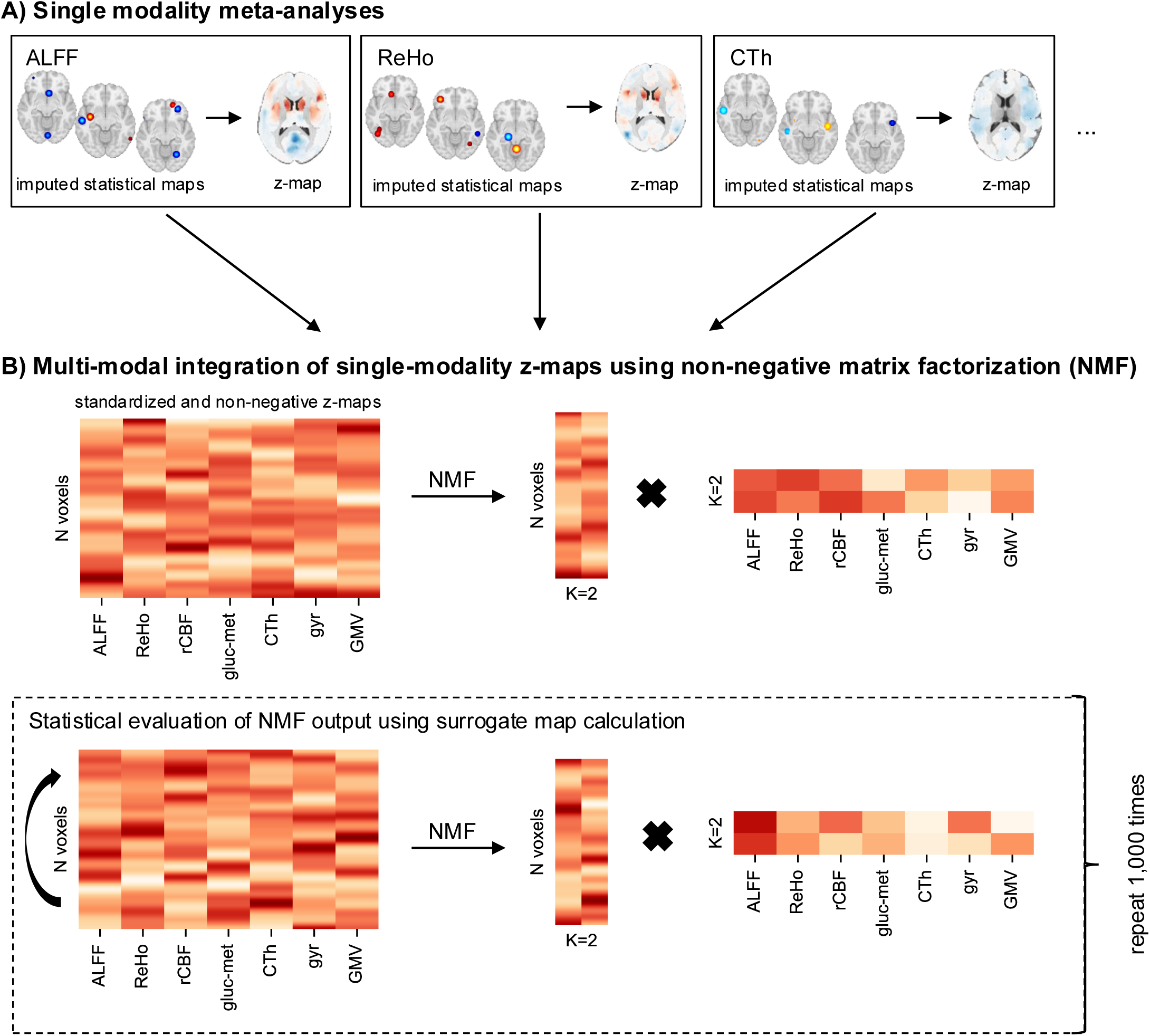
Overview of single-modality meta-analyses and multimodal integration. **(A)** For each of the seven investigated modalities (i.e., amplitude of low-frequency fluctuations (ALFF), regional homogeneity (ReHo), regional cerebral blood flow (rCBF), glucose metabolism (gluc-met) based on [^18^F]-Fluorodeoxyglucose positron emission tomography, cortical thickness (CTh), gyrification (gyr), and gray matter volume (GMV) based on voxel-based morphometry with modulation), we conducted a single-modality meta-analysis using the Seed-based *d* Mapping with Permutation of Subject Images (SDM-PSI) software package (https://www.sdmproject.com/software/; (47)). In brief, voxel-wise effect-size maps are imputed by the information of reported peak coordinates and their statistics. Based on these imputed maps for each original study, maps of means across studies are computed using a random-effects model (z-maps). **(B)** The z-maps resulting from the seven single-modality meta-analyses were integrated using non-negative matrix factorization (NMF). For this, z-maps were first standardized and shifted to non-negativity (matrix on the left). Then, the N voxel x 7 modalities matrix was decomposed into a component matrix W (N voxels x 2 components) and a weight matrix H (2 components x 7 modalities; see Supplementary Methods for a detailed description). To assess statistical significance of NMF-derived component maps, the decomposition using NMF was conducted 1,000 times, each time with a different input matrix. For this, the original input matrix was shuffled while preserving spatial autocorrelation in each input modality (52). Thereby, a voxel-wise null distribution is generated, which can be used to compute empirical p-values.

To identify clusters in the component maps, FSL’s function *cluster* with threshold 0.0075 was applied. To further characterize the resulting clusters, we quantified their overlap with the anatomical regions of the Harvard-Oxford atlas derived from FSL (53–56) and the seven canonical resting-state networks identified by Yeo and colleagues (57) using a custom Python script.

### Linking convergent multimodal macroscopic brain abnormalities with cell-type expression

To investigate whether consistent multimodal abnormalities in schizophrenia preferentially localize to regions abundant with specific cell types, we tested the spatial overlap of significant OPNMF-derived components with cell-type specific gene expression patterns derived from the Allen Human Brain Atlas (AHBA) (58,59) and single-cell expression studies (60–64) as presented in previous work (44,65).

Direct quantification of cellular abundance in vivo would currently require human brain tissue samples. Therefore, to approximate regional cellular distributions in the human brain, previous studies (44,65) relied on data from the AHBA (https://human.brain-map.org/; Supplementary Methods) and multiple single-cell studies of the adult human cortex (60–64) to derive a set of cell type-specific gene markers for the adult human brain. These markers cover the following cell types: astrocytes, endothelial cells, microglia, oligodendrocyte progenitor cells (OPCs), oligodendrocytes, excitatory neurons, and inhibitory neurons. To generate regional representations of cellular abundance in the cortex, mean regional expression scores were computed for each cell type using the preprocessed AHBA gene expression data.

To spatially correlate convergent multimodal macroscopic brain abnormalities with cell type-specific gene expression, we first transformed the significant OPNMF component from MNI volumetric space to *fsaverage5* surface space and parcellated it into 68 cortical regions with the Desikan-Killiany atlas (56) using the *neuromaps* toolbox (version 0.0.5) (66). Next, also using *neuromaps*, we conducted spatial autocorrelation-preserving permutation tests (“spin tests”, 1000 iterations), which have been proposed (among other methods) to evaluate the statistical significance of the spatial overlap between two brain maps (66–68) (for details, see Supplementary Methods).

### Linking convergent multi-modal macroscopic brain abnormalities with recent evolutionary change

To examine whether convergent multi-modal abnormalities primarily overlap with cortices under recent evolutionary change in humans, we used previously compiled data sources of human-specific connectivity (39,69) and human-specific cortical expansion (34), both compared to chimpanzees (see Supplementary Methods for details about the generation of these maps). We investigated both measures since they represent distinct aspects of evolutionary reorganization (70). Spatial overlap was assessed using permutation-based Dice coefficients or spatial correlations with appropriate spatial null models, respectively. An independent human-specific connectivity map served as sensitivity analysis. For details, see Supplementary Methods.

## Results

### Single-modality coordinate-based meta-analyses of macroscopic brain abnormalities in schizophrenia

The literature search yielded a total of 209 studies from 200 publications (as 9 publications investigated more than one imaging method), comprising 11,794 patients with schizophrenia and 10,342 healthy controls and encompassing 28 ALFF, 25 ReHo, 14 rCBF, 4 glucose metabolism, 21 CTh, 11 gyrification, and 106 GMV experiments. Supplementary Results and Supplementary Table S1 provide demographical information of all included studies.

Regarding functional measures, ALFF was higher in patients with schizophrenia compared to controls in the thalamus and inferior frontal gyrus (results reported for p<0.001; Fig. 2; see Supplementary Table S2 for MNI peak coordinates of significant clusters), and lower in the postcentral gyri, corpus callosum, and left cerebellum. ReHo was higher in the bilateral striatum, bilateral superior frontal regions, left insula, and left inferior temporal gyrus in patients compared to controls. We identified lower ReHo in bilateral inferior frontal, occipital, and the postcentral cortices as well as right precuneus and right inferior temporal gyrus. rCBF was higher in the left lenticular nucleus of the putamen and lower in left frontal, left anterior cingulate, and right middle occipital regions. Glucose metabolism was increased in the right inferior temporal gyrus and decreased in the left superior frontal gyrus.

**Figure 2:**
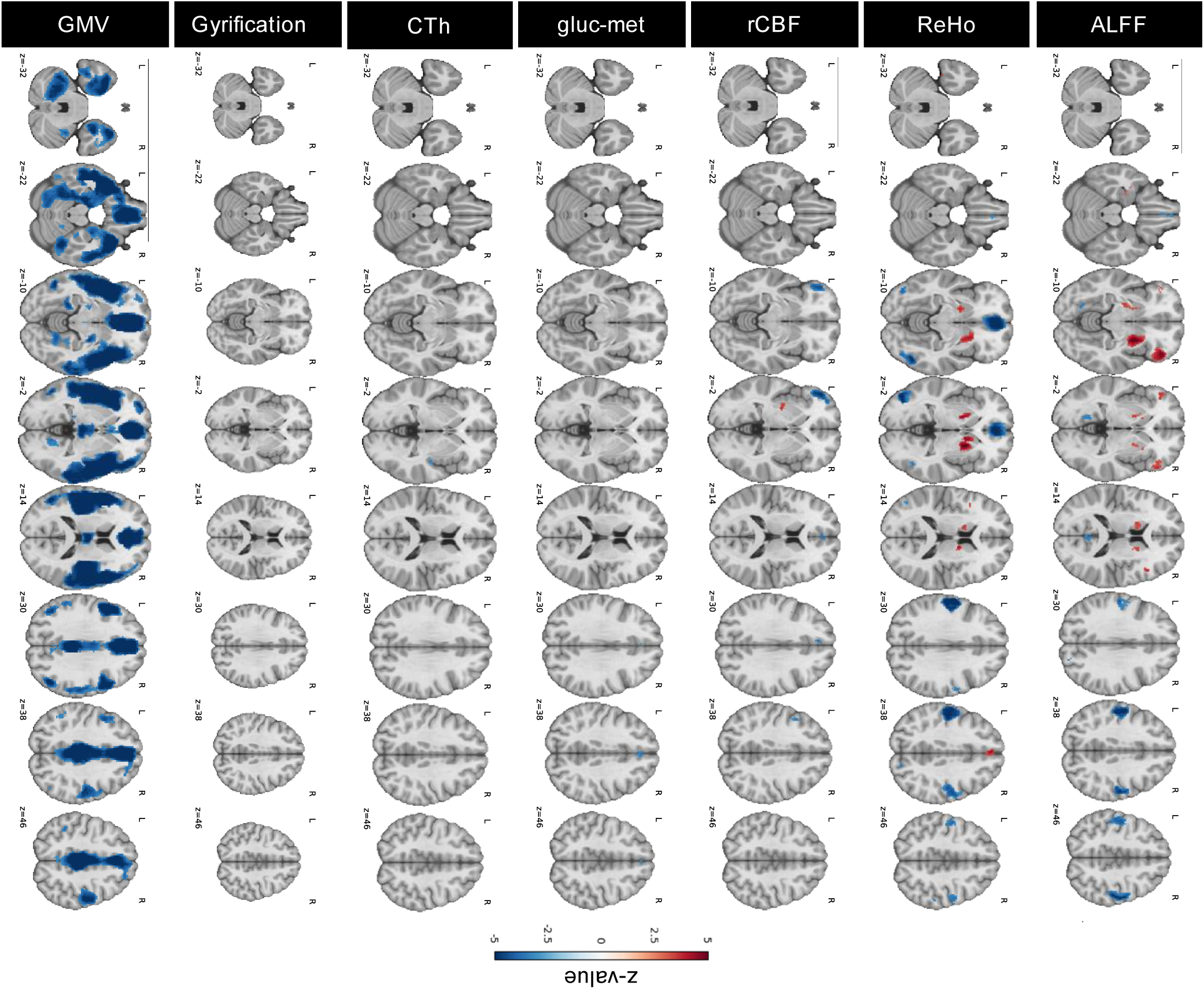
Single-modality meta-analyses results. Outcome maps of the seven meta-analyses for each modality, representing the difference between patients with schizophrenia and healthy controls presented as z-values. Investigated modalities included amplitude of low-frequency fluctuations (ALFF), regional homogeneity (ReHo), regional cerebral blood flow (rCBF), glucose metabolism (gluc-met) based on [^18^F]-Fluorodeoxyglucose positron emission tomography, cortical thickness (CTh), gyrification, and gray matter volume (GMV) based on voxel-based morphometry with modulation. Results are presented as z-values, thresholded for significance (p<0.001). Unthresholded maps are presented in Supplementary Figure S2.

Regarding structural measures, CTh was reduced in the right insular cortex. No significant alterations in gyrification were detected. Patients exhibited widespread lower VBM-derived GMV in bilateral temporal and mediofrontal areas, left thalamus, and left hippocampus. Across modalities, most peaks were not driven by significant inter-study heterogeneity or publication bias (Supplementary Results, Supplementary Table S3), indicating reliability of the results.

### Integrated multimodal macroscopic brain abnormalities converge on insular, mediofrontal, and sensorimotor cortices

Integration of the seven meta-analytic results maps with OPNMF (K=2) and thresholding for statistical significance using spatial autocorrelation-informed null models identified a single significant multimodal convergence pattern centered in bilateral insular, mediofrontal, and sensorimotor regions (Fig. 3). The component comprised multiple clusters extending to the temporal pole and orbitofrontal cortex, together with medial frontal clusters (Fig. 3C, Supplementary Table S4): the first cluster (cluster index 19, 10358 voxels) spanned the right frontal pole, insula, orbitofrontal cortex, and the central opercular cortex. The second cluster (cluster index 18, 8120 voxels) included the left insula, temporal pole, superior temporal gyrus, and the central opercular cortex. When comparing the spatial outline of these regions with well-established resting-state connectivity networks (57), we found a large overlap with the default mode, salience, and somatomotor networks. A further cluster (cluster index 17, 1483 voxels) spanned bilateral superior frontal, paracingulate, and cingulate gyri, which lie in the frontoparietal control and salience networks. Further smaller clusters spanned the posterior cingulate cortices and frontal areas, corresponding to parts of the default mode and salience network. For a detailed description of cluster sizes, MNI peak coordinates, and network overlap, see Supplementary Tables S4 and S5. The strongest NMF loadings were observed for ALFF, rCBF, and ReHo (Fig. 3B), indicating spatial co-localization of functional activity, perfusion, and local synchronization alterations.

**Figure 3:**
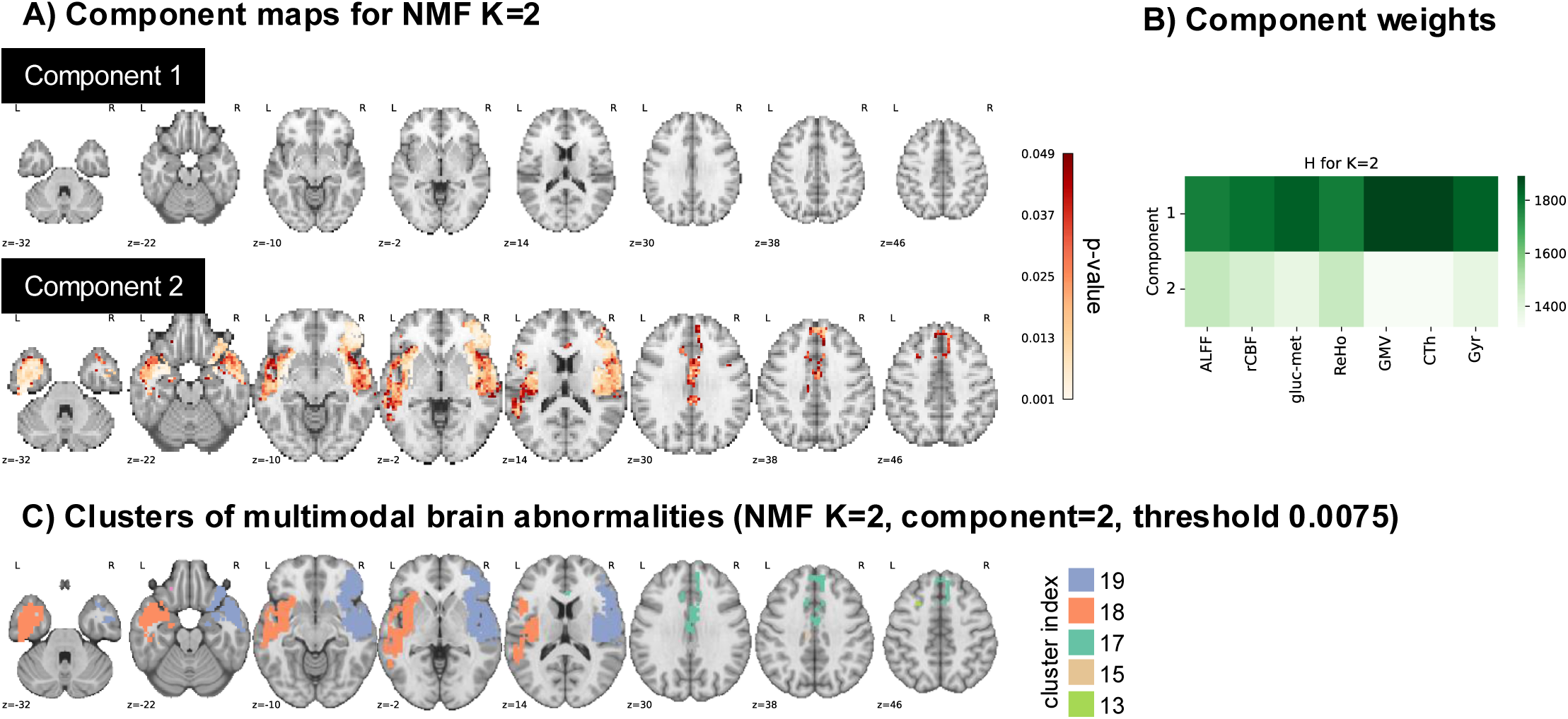
Multi-modal abnormalities in schizophrenia converge on insular, mediofrontal, and sensorimotor cortices. Results of the multimodal integration using non-negative matrix factorization with two components (K=2). **(A)** Spatial maps of the resulting components 1 and 2. Only significant voxels are depicted (p_SMASH_<0.05). **(B)** Component weights for each imaging modality: amplitude of low-frequency fluctuations (ALFF), regional homogeneity (ReHo), regional cerebral blood flow (rCBF), glucose metabolism (gluc-met) based on [^18^F]-Fluorodeoxyglucose positron emission tomography, cortical thickness (CTh), gyrification (gyr), and gray matter volume (GMV) based on voxel-based morphometry with modulation. **(C)** Clusters of NMF results (K=2, component=2) identified with FSL cluster using a threshold of 0.0075. Refer to Supplementary Table S4 for peak coordinates, the number of voxels per cluster, and anatomical locations.

Sensitivity analyses confirmed the robustness of this spatial pattern. Increasing the number of components (K = 3) produced a consistent spatial pattern that was nearly identical with the K=2 solution: the spatial Spearman correlation coefficient between component 2 of K=2 and component 2 of K=3 was Spearman’s ρ=0.979 (Supplementary Results, Supplementary Figure S4). Furthermore, to investigate an alternative multimodal integration strategy, we substituted OPNMF with PCA. To yield results which are comparable with the OPNMF framework, two input matrix preprocessing strategies were compared to ensure non-negativity: using the absolute value of all entries (PCA-abs) and modality-wise z-score normalization followed by subtraction of the global minimum value (PCA-min; Supplementary Methods). Spatial correspondence between the first principal components of both strategies and component 2 from OPNMF was very high (Spearman’s ρ = −0.950 for PCA-min; Spearman’s ρ = 0.735 for PCA-abs; see Supplementary Results, Supplementary Figure S5). Together, these analyses indicate that the identified insular-mediofrontal-sensorimotor pattern is stable across decomposition parameters and integration approaches.

### Multimodal brain aberrations in schizophrenia are associated with glial gene expression profiles

To assess the cell-biological underpinnings of convergent multimodal abnormalities in schizophrenia, we tested for spatial correspondence between the significant OPNMF component (component 2), and the cortical distribution of eight major brain cell types, approximated by gene expression derived from the AHBA (58).

To achieve this, the component scores were mapped onto cortical surfaces and parcellated into 68 Desikan–Killiany regions (56). Significant spatial correspondence was observed for the gene expression profiles of astrocytes (Spearman’s ρ=0.471, p_spin_=0.023), microglia (ρ=0.430, p_spin_=0.035), and OPCs (ρ=0.549, p_spin_=0.006) but not of other cell types (Fig. 4; Supplementary Table S6).

**Figure 4:**
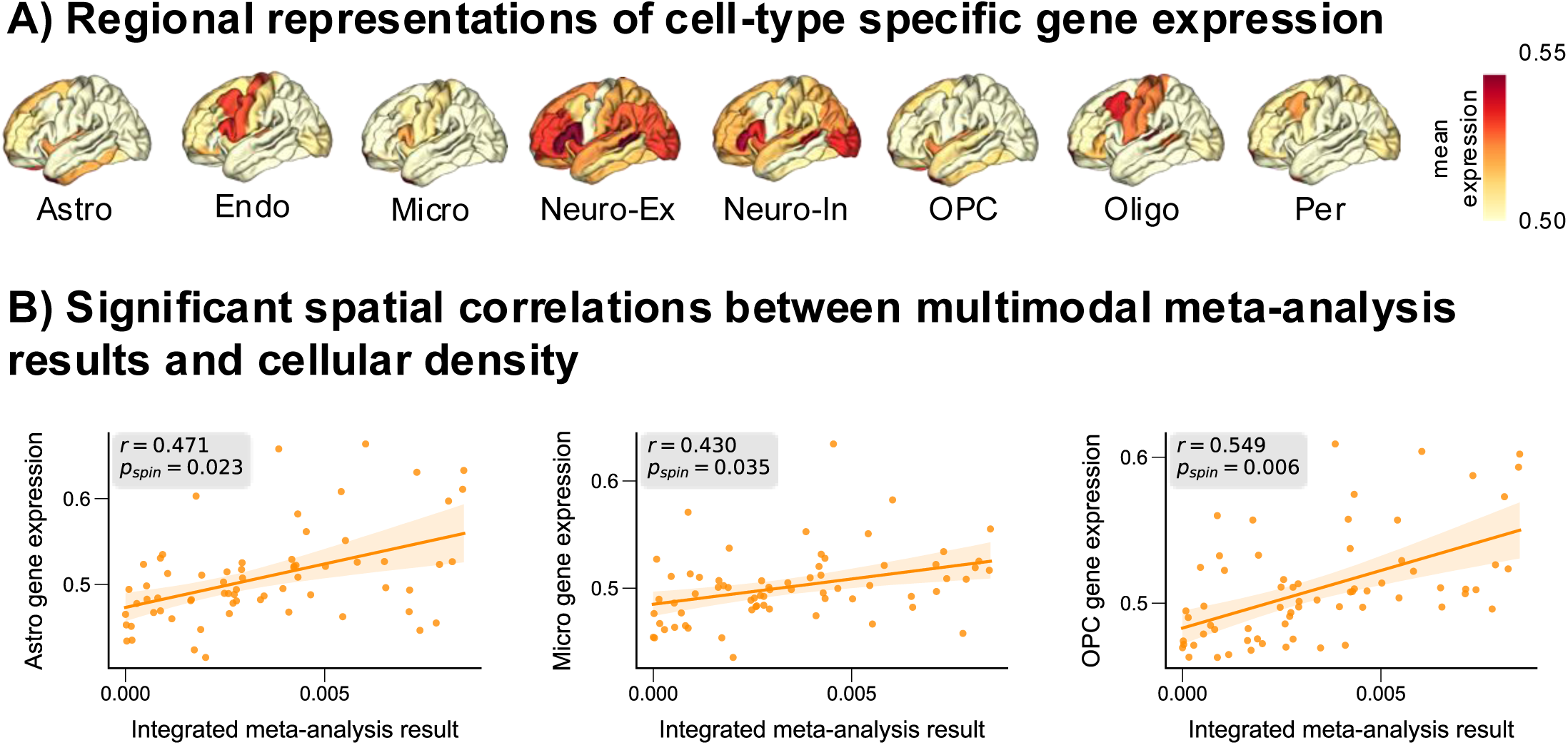
Multimodal brain abnormalities in schizophrenia are related to cell-type specific gene expression of astrocytes, microglia, and oligodendrocyte precursor cells. Mean expression profile of eight brain cell types were based on sets of cell type-specific marker genes derived from single-cell RNA studies (60–64) as well as regional gene expression information from the AHBA (58). The investigated cell types comprised astrocytes (Astro), endothelial cells (Endo), microglia (Micro), excitatory neurons (Neuro-Ex), inhibitory neurons (Neuro-In), oligodendrocytes (Oligo), oligodendrocyte precursor cells (OPC), and pericytes (Per).

### No evidence for preferential localization of multimodal brain abnormalities to cortices with recent evolutionary change

To test whether convergent multimodal abnormalities in schizophrenia preferentially localize to cortices with recent evolutionary change in humans relative to chimpanzees, we assessed the spatial overlap between the significant OPNMF component (component 2; K=2) and human-specific cortical signatures compared to chimpanzees derived from two complementary but distinct approaches: (i) cortical representation of reorganized white matter connectivity and (ii) cortical expansion, which represents the disproportionate enlargement of cortices in the human lineage thought to reflect the addition of novel cortical columns through a prolonged period of neuronal progenitor expansion (34,70) (Fig. 5).

i. The overlap with the cortical representation of human-specific connectivity was low and not significant (Dice coefficient=0.368; p_perm_=0.483). Consistent results were observed when using spatial Spearman correlation instead of the Dice coefficient (Spearman’s ρ=-0.328, p_SMASH_=0.455). For a further control analysis, we correlated the multimodal brain abnormalities with another human-specific connectivity difference map, reflecting the divergence of the overall connectivity profile between humans and chimpanzees (69). Likewise, there was no significant association between the two maps (Spearman’s ρ=0.210, p_spin_=0.146; Supplementary Methods, Supplementary Figure S6).
ii. No significant relationship was observed between the multimodal abnormalities in schizophrenia and cortical expansion (Spearman’s ρ=0.039, p_spin_=0.879).

**Figure 5:**
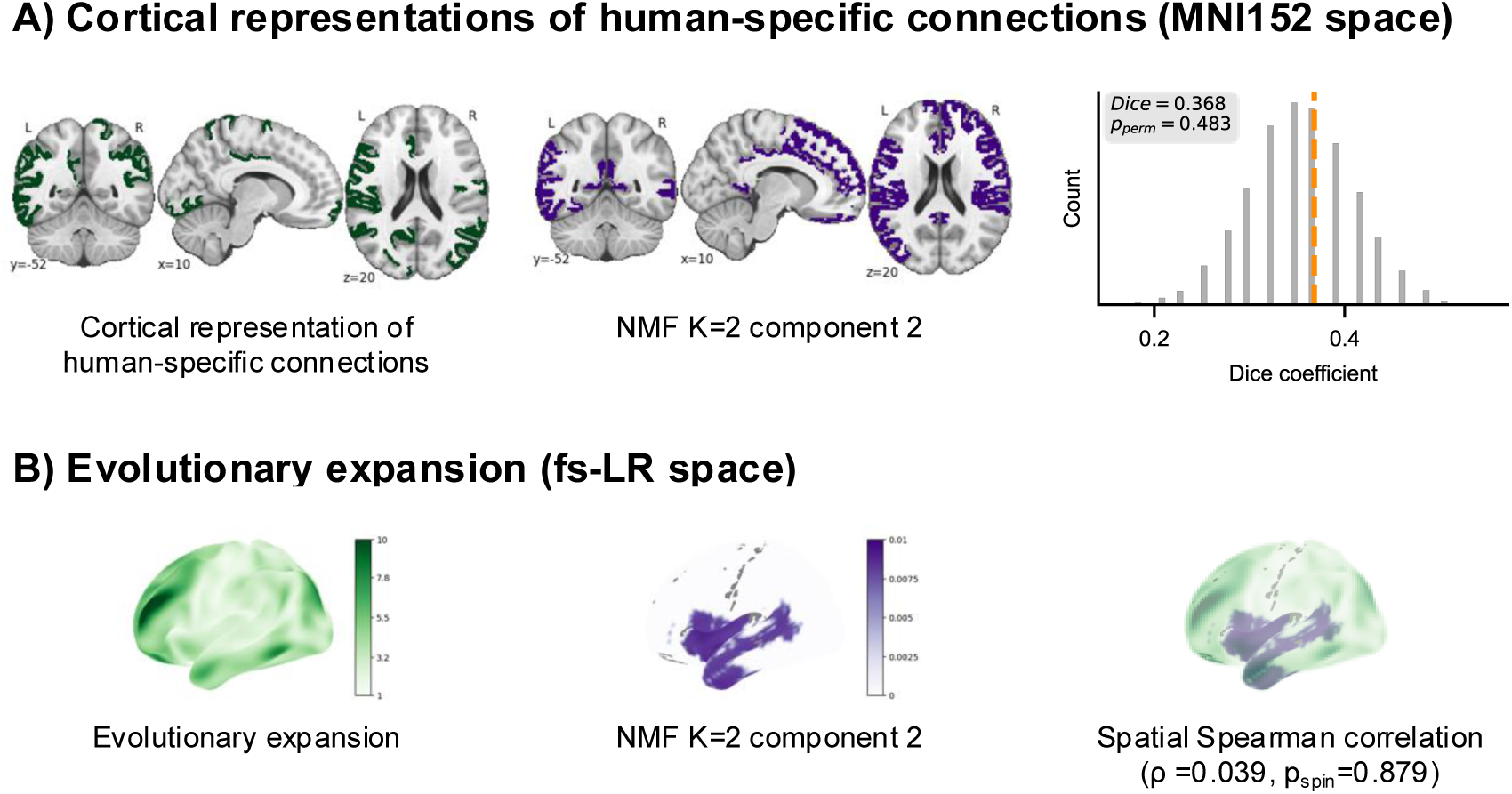
Multimodal brain abnormalities in schizophrenia are not restricted to evolutionarily novel brain regions. Maps representing evolutionary novelty were derived from previous work (left image in each line), using either the cortical representation of white matter connectivity differences between humans and chimpanzees **(A)** or the emergence of novel cortical columns (i.e., cortical expansion) from humans to chimpanzees **(B)** (34,39). The output of the integrated meta-analysis was resampled to the respective space in which the evolutionary novelty map was available (middle image in each line). The spatial overlap was then quantified as the Dice coefficient of binarized maps (for A) or as the spatial Spearman correlation coefficient (B).

In summary, we found no evidence that converging multimodal schizophrenia-related brain abnormalities are preferentially localized to cortices under recent evolutionary change, at least regarding the investigated modalities and maps.

## Discussion

To test for multimodal convergence of macroscopic brain abnormalities in schizophrenia and their biological underpinnings in terms of cell types and evolutionary novelty, we conducted seven single-modality coordinate-based meta-analyses of in total 200 neuroimaging studies, comprising over 11,000 patients with schizophrenia and 10,000 healthy controls. By integrating these results using OPNMF, we identified a multimodal spatial convergence pattern focusing on insular, mediofrontal, and sensorimotor cortices, which was robust across decomposition methods and component numbers. This convergence pattern was spatially aligned with “glia-rich” (based on the abundance of marker gene expression) cortical regions but not with cortices of recent evolutionarily change relative to chimpanzees and with respect to connectivity and cortical expansion. These results support glia-related regional vulnerability as an organizing principle of multimodal brain abnormalities in schizophrenia.

### Multimodal macroscopic brain abnormalities in schizophrenia converge on insular, mediofrontal, and sensorimotor regions

OPNMF-based multimodal integration of meta-analytically significant abnormalities in patients with schizophrenia regarding CTh, gyrification, VBM-derived GMV, ALFF, ReHO, rCBF, and glucose metabolism revealed a spatial convergence pattern centered in insular, mediofrontal, and sensorimotor cortices, overlapping mostly with default-mode, salience, and sensorimotor resting-state networks. Control analyses suggested that the results were robust regarding the integration method and not significantly biased by heterogeneity or publication bias.

Our results are in line with previous unimodal and multimodal meta-analyses, which also identified insular, frontal, and sensorimotor regions as the major involved regions as well as default-mode, salience, and sensorimotor networks as major relevant networks (21,71,72), supporting the reliability of our approach. Furthermore, we extend previous work (21) by incorporating additional imaging modalities and using a different integration method, namely OPNMF, which identifies latent covariance patterns rather than simple spatial overlap.

Attempting a careful functional and pathophysiological interpretation of the regional focus of our findings, the insula has been proposed as a central hub of the salience network, responsible for mapping internal events and determining which external stimuli the brain pays attention to (73–75), a function often disturbed in patients with schizophrenia. Aberrant anatomy of the insula, for example decreased GMV, has been repeatedly identified in schizophrenia and prodromal stages (76,77). Medio-frontal regions, including the paracingulate and cingulate gyri, are hypothesized as important components of the default-mode network and are thought to be essential for integrating cognitive and emotional processing (2,78), which is frequently impaired in schizophrenia. Their involvement across diverse modalities (21,79,80) suggests a stable pathological signature disrupting the functional integration particularly associated with complex social behaviors. Sensorimotor regions and networks are frequently implicated in pathophysiological theories of schizophrenia, e.g., in the context of hallucinations (81,82). Meta-analytic work has repeatedly shown reduced local functional connectivity or spontaneous activity in the postcentral and precentral gyri, as well as schizophrenia-specific convergence of dysconnectivity and gray-matter loss in the lateral postcentral cortex (83,84). Finally, default-mode, salience, and sensorimotor networks are located along a principal gradient of fundamental macroscopic brain organization from unimodal to transmodal cortices (85).

Taken together, the multimodal convergence on insular, mediofrontal, and sensorimotor cortices supports a model in which schizophrenia involves coordinated disruption of medial frontal regulatory regions, salience network hubs, and systems supporting bodily and motor representations.

### Multimodal abnormalities preferentially occur in regions with high glial abundance but do not appear restricted to cortices under recent evolutionary change

The converging multimodal abnormalities preferentially occurred in cortical regions with high glial abundance based on the abundance of marker gene expression. This supports theories that suggest changes of neuron-glia interactions in schizophrenia pathophysiology, probably in addition to neuronal dysfunction (5,28,29). Although large-scale GWAS consistently show that inherited risk is concentrated in genes expressed in excitatory and inhibitory neurons (7,30), our results suggest that the identified multimodal abnormalities spatially align more closely with gene expression patterns of astrocytes, microglia, and OPCs. Our data does not permit further speculation about the lack of overlap with neuronal gene expression patterns. We want to stress the methodological differences between “non-spatial” GWAS approaches, which identify risk genes for schizophrenia in the genome, and the regionally specific gene expression maps employed here, with which we assessed a spatial correspondence between regional gene expression and convergent multimodal brain aberrations. Together, these observations may suggest that neuronal genetic risk and glial vulnerability may represent complementary rather than competing levels of disease biology (28).

Our findings are consistent with the hypothesis that early microglial activation during embryogenesis disrupts the differentiation and maturation of glial progenitor cells, leading to delayed oligodendrocyte and astrocyte development and, ultimately, to the hypomyelination and impaired synaptic support observed in schizophrenia (5,28,86). Experimental evidence from humanized chimeric mice also supports this model, demonstrating that schizophrenia-derived OPCs exhibited impaired maturation leading to reduced white matter integrity and behavioral deficits (86). Likewise, postmortem studies report reduced numbers of oligodendrocytes and astrocytes in schizophrenia (28,29,87). Against this background, our results suggest that brain regions with a higher abundance of glial cells may be particularly vulnerable to the biological processes underlying schizophrenia, resulting in convergent abnormalities across multiple imaging modalities.

An important consideration for this interpretation is the marked biological heterogeneity of schizophrenia. Rather than representing a single disease entity, schizophrenia is increasingly viewed as a syndrome in which multiple biological pathways converge on overlapping but still heterogeneous clinical phenotypes (88,89). Recent normative modelling studies demonstrated that this heterogeneity manifests in brain anatomy (15,16,31) and that cellular mechanisms may underlie different neuroanatomical subtypes, with one patient subgroup exhibiting cortical thinning predominantly associated with neuronal gene expression, while a larger subgroup showed thinning in regions enriched for astrocytes, microglia, and OPCs (31). Despite this heterogeneity across individuals and imaging modalities, the present findings demonstrate that these diverse abnormalities converge on a reproducible set of cortical regions. This suggests that heterogeneity at the level of individual neuroimaging measures does not exclude the existence of common organizational principles at the systems level. Identifying such principles may provide a more stable framework for linking macroscopic imaging findings to underlying cellular mechanisms. Future studies combining multimodal neuroimaging with transcriptomics and genetics at the individual level will be needed to test this hypothesis directly.

Contrary to our hypothesis, multimodal abnormalities in schizophrenia were not preferentially localized to cortices with recent evolutionary change. Potential reasons for this null finding might be: first, we investigated the spatial correlation between multimodal neuroimaging abnormalities and maps of evolutionary reorganization; partial or network-level overlap may not be captured by this method. Second, previous studies implicating human brain evolution in schizophrenia have primarily focused on specific aspects of brain organization; for example, van den Heuvel and colleagues demonstrated that schizophrenia disproportionately affects human-specific structural connections (39), an effect that may not hold across the broader range of imaging modalities examined in the present study. Slight methodological differences between our approach and other studies (e.g., we focused on regions instead of tracts) might also contribute. Third, evolutionary aspects may be more relevant at the level of genetic risk than regarding the spatial distribution of macroscopic brain pathology. Taken together, our results do not contradict previous evidence and theories linking schizophrenia to human brain evolution. Rather, they must be strictly interpreted in the specific context of the current study and the applied methods. Future studies should integrate evolutionary genomics with multimodal neuroimaging to address these issues.

### Limitations

First, we conducted coordinate-based rather than image-based meta-analyses. Because coordinate-based approaches use reported peak coordinates and associated effect sizes rather than unthresholded statistical maps, they could discard spatial information and be less sensitive (90,91). Nevertheless, because whole-brain statistical maps remain rarely available, coordinate-based meta-analysis represents a viable approach for large-scale analyses.

Second, the number of available studies differed considerably across imaging modalities, with VBM contributing substantially more studies compared to other modalities, while only a limited number of FDG-PET studies met the inclusion criteria. Although OPNMF yielded robust decomposition across multiple sensitivity analyses, this imbalance may still have influenced the resulting components. Furthermore, as OPNMF is an unsupervised data-driven approach, the identified components represent latent covariance patterns specific to the present data and generalizability might therefore be limited.

Third, the included studies varied considerably in clinical and methodological characteristics, including medication status, illness duration, symptom severity as well as image acquisition and preprocessing, all of which may have contributed to between-study heterogeneity, although we controlled for that. Fourth, the analysis of spatial correspondence relied on cortical cell-type maps derived from healthy controls and thus assumes that large-scale regional expression patterns are sufficiently conserved across individuals and remain broadly stable in disease. Although these assumptions have been justified previously (44,59), non-invasive tools to investigate this are lacking. Crucially, spatial correspondence analyses cannot establish causal relationships or identify the cellular mechanisms responsible for neuroimaging abnormalities. Instead, they provide a framework for generating biologically informed hypotheses regarding the cellular architecture that may shape regional vulnerability to schizophrenia-related brain abnormalities.

### Conclusion

By integrating evidence across multiple structural and functional neuroimaging modalities, we identified patterns of convergent macroscopic brain abnormalities in schizophrenia that were related to glial gene expression profiles, providing further support for emerging models of glial dysfunction as a feature of schizophrenia pathophysiology (5,28). These findings establish a multiscale framework linking convergent neuroimaging abnormalities with regional cellular architecture and provide a foundation for dissecting the biological heterogeneity of schizophrenia.

## Data Availability

All data produced in the present study are available upon reasonable request to the authors. Analysis code is publicly available at https://github.com/Melissa1909/schizophrenia-multimodal.

## Code availability

All code used to perform the analyses presented in this paper can be found at https://github.com/Melissa1909/schizophrenia-multimodal.

## Acknowledgements and disclosures

M.T. was supported by the German Center for Mental Health (Deutsches Zentrum für Psychische Gesundheit DZPG). F.B. was supported by the Else Kröner Memorial Stipendium, German Research Foundation (Deutsche Forschungsgemeinschaft DFG), and German Center for Mental Health (Deutsches Zentrum für Psychische Gesundheit DZPG). C.S. was supported by the German Research Foundation (DFG), C.M. was supported by the National Natural Science Foundation of China and Natural Science Foundation of Sichuan Province. J.P. was supported by the German Center for Mental Health (Deutsches Zentrum für Psychische Gesundheit DZPG, FKZ.: 01EE2303B). The authors report no conflicts of interest.

