## Supplementary Material for "Multimodal macroscopic brain abnormalities in schizophrenia converge on insular, mediofrontal, and sensorimotor cortices and link with glial cell expression but not with recent evolutionary change"

for

by

Thalhammer et al.

Spatial overlap analysis between human-specific cortical expansion and multi-modal brain abnormalities 13

### Supplementary Methods

#### Coordinate-based meta-analysis

##### *Systematic literature search*

We conducted a systematic literature search on PubMed for seven structural and functional modalities, namely cortical thickness (CTh), gyrification, voxel-based morphometry (VBM) with modulation interpreted as gray matter volume (GMV), amplitude of low-frequency fluctuations (ALFF), regional homogeneity (ReHo), arterial spin labelling-derived regional cerebral blood flow (rCBF), and glucose metabolism based on [$^{18}$F]-Fluorodeoxyglucose positron emission tomography (FDG-PET). The following search queries were used for the respective modality:

- ALFF: schizo*[Title/Abstract] AND (“ALFF”[Title/Abstract] OR “amplitude of low-frequency oscillations”[Title/Abstract] OR “amplitude of low frequency fluctuations”[Title/Abstract])
- ReHo: schizo*[Title/Abstract] AND (“ReHo”[Title/Abstract] OR “regional homogeneity”[Title/Abstract])
- rCBF: schizo*[Title/Abstract] AND ("cerebral blood flow"[Title/Abstract] OR "arterial spin labeling"[Title/Abstract] OR "arterial spin labelling"[Title/Abstract] OR (perfusion [Title/Abstract] AND (MRI[Title/Abstract] OR "magnetic resonance imaging"[Title/Abstract])))
- FDG-PET: schizo*[Title/Abstract] AND (“cerebral glucose metabolism"[Title/Abstract] OR “FDG”[Title/Abstract])
- Cortical thickness: schizo*[Title/Abstract] AND "cortical thickness"[Title/Abstract]
- Gyrification: schizo*[Title/Abstract] AND "gyrification"[Title/Abstract]
- VBM: schizo*[Title/Abstract] AND ("VBM"[Title/Abstract] OR "voxel-based morphometry"[Title/Abstract])

This was supplemented by hand-searching of reference lists in review articles or the reference list of other studies.

##### *Study selection*

Magnetic resonance imaging (MRI) and [$^{18}$F]-Fluorodeoxyglucose positron emission tomography (FDG-PET) studies were included in the meta-analysis if they satisfied the following criteria: (i) the study was original (rather than a review or abstract, etc.) and published in an English language peer-reviewed journal; (ii) the study compared the respective imaging modality between patients formally diagnosed with schizophrenia according to DSM, ICD, or similar criteria and healthy controls; (iii) the respective modality of interest was analyzed at the whole-brain level; (iv) peak coordinates were reported in three-dimensional stereotactic coordinates (i.e., Talairach or Monreal Neurological Institute (MNI)); (v) if the study was longitudinal, only baseline data were included for analysis.

Exclusion criteria were: (i) the study was published in another language than English; (ii) the full article could not be accessed; (iii) there was no explicit diagnosis of schizophrenia; (iv) the analysis examined regions-of-interest only instead of the whole brain; (v) three-dimensional stereotactic coordinates of the peak of activation point were unavailable or not reported in stereotactic space (i.e., Talairach or MNI); (vi) less than 10 participants in a single group were examined; (vii) if baseline data of a longitudinal investigation were unavailable.
There were no restrictions regarding participant age, disease duration, symptom severity (current episode or remission), or medication status to ensure maximum study coverage. Refer to Supplementary Figures S1 for an overview of the process for each modality.

*Coordinate-based meta-analyses using SDM-PSI*

For each imaging modality, we conducted a separate meta-analysis using Seed-based *d* Mapping with Permutation of Subject Images (SDM-PSI; version 6.23; https://www.sdmproject.com/software/; (1)) on a Debian GNU/Linux 11 (bullseye) running a Linux kernel 5.10.0-35-amd64. SDM-PSI uses the reported peak coordinates and their statistics to recreate voxel-wise effect size maps of the difference between patients and controls for each original study using an anisotropic non-normalized Gaussian kernel (20 mm Full Width at Half Minimum) and a voxel size of 2 mm isotropic (2). For surface-based modalities (i.e., cortical thickness and gyrification), a correlation template restricted to cortical gray matter ("FreeSurfer mask" instead of "gray matter") was used. Next, maps of means across studies were computed using a random-effects model, weighted by sample size, intra-study variability, and between-study heterogeneity. Multiple imputations were weighted according to Rubin's rules (1). The default settings of the SDM-PSI software were used (i.e., peak height $Z > 1$; $p < 0.005$, uncorrected; cluster extent > 10 voxels, 50 imputations).

*Data extraction and quality assessment*

Study quality was systematically assessed in terms of data accuracy, study design, and demographic, and clinical characteristics. To control for potential confounding factors, publications with identical patient and healthy control (HC) samples were treated as a single study. If a study compared multiple patient samples (e.g., hallucinating patients vs. HC and non-hallucinating patients vs. HC) with the same control sample, only the analysis of the main effect was considered (e.g., HC vs. hallucinating and non-hallucinating patients).
As a first step, the abstracts of the studies found through the keyword search in the PubMed database were screened for the inclusion criteria. If these were met, the name of the first author, year of publication, and the publication title were noted in an Excel sheet.
Secondly, the Methods section of the paper was reviewed for the examined contrast between patients with schizophrenia and healthy controls for the respective modality as well as for the number of investigated participants per group. If all inclusion criteria applied, the number of investigated

participants per group, the reported peak coordinates, the stereotactic space (i.e., Talairach or MNI), the software the coordinates were extracted with (i.e., “SPM”, “FSL”, or “other”), the t-value statistical threshold, and the direction of the observed effect (i.e., less/more) were noted down. If no coordinate-based whole-brain results were reported, the study was excluded. Furthermore, if available, the respective z-statistic, t-statistic, and p-values were extracted. Z-values and p-values were converted into T-values using the online converter provided by the SDM-PSI toolbox (https://www.sdmproject.com/utilities/?show=Statistics), considering the number of included covariates to calculate the degrees of freedom. If studies reported non-significant results, peak coordinates, sample sizes, as well as respective statistics were recorded in a separate Excel file. T-value thresholds of statistical significance were set to 3.1 if unsure or not available for a particular study as recommended as a conservative threshold by the SDM-PSI tutorial (https://www.sdmproject.com/software/tutorial.pdf, version January 2019).

The information gathered in two Excel files for each modality (i.e., studies reporting significant and non-significant results) were used as an input for a MATLAB script, which extracts all relevant information and writes them into one .txt file per study. These .txt files will then be used as an input for subsequent meta-analysis using the SDM-PSI toolbox (for a detailed explanation of the required input, see the tutorial of SDM-PSI (https://www.sdmproject.com/software/tutorial.pdf, version January 2019) and (3)). The file name encodes the first author’s name, the software the peak coordinates were extracted with, and the stereotactic space of reported peak coordinates, and contains the coordinates and t-value of each peak in a different row (3).

In addition, demographic and clinical information were extracted from each study. This included sample sizes and mean age for both groups, as well as illness duration, information about the medication status, and the scores on the Positive and Negative Syndrome Scale (PANSS) for patients if available.

*Assessment of heterogeneity and potential publication bias*

To examine heterogeneity and potential publication bias, peak coordinate values for each significant meta-analytic peak were extracted. Between-study heterogeneity was assessed using the $I^2$-statistic, which quantifies the overall variation in each set of results that is not attributable to chance, but rather to differences between studies. Values below 25% indicate low heterogeneity and greater robustness of the findings, whereas values below 50% suggest medium heterogeneity (4). Additionally, to determine possible publication bias, funnel plots were generated for visual inspection, and Egger tests were conducted (5,6). Significant publication bias was inferred when the funnel plot appeared asymmetric, and the Egger test p-value was below 0.05.

#### Multi-modal integration

*Non-negative matrix factorization (NMF) background*

To examine spatial convergence of brain aberrations in schizophrenia, we applied non-negative matrix factorization (NMF) on the unthresholded z-maps of the single-modality meta-analyses. NMF has been shown to produce parts-based representations of the brain that are highly interpretable and biologically plausible and are therefore well suited for neuroimaging data, for example to study covariance across multiple measures (7–9).

In brief, NMF decomposes an input matrix V of dimensions (m x n) into a component matrix W (m x k) and a weight matrix H (k x n). Thereby, k is the number of pre-defined components, m is the number of imaging modalities, and n is the number of non-NaN voxels. The aim is to decompose the input matrix V so that the multiplication of W and H reconstructs the original input as accurately as possible by minimizing the reconstruction error between the original input V and the reconstruction output (W x H) (7,10,11) (Supplementary Figure S3).

Thereby, a non-negativity constraint is imposed on all elements of W and H. Because neither matrix may contain negative values, the reconstruction of V is purely additive: each voxel's signal profile is expressed as a weighted sum of components, where weights reflect the degree to which each component contributes at that location. No component can cancel out another, which encourages the algorithm to discover a parts-based representation of the data, i.e., a set of components that correspond to distinct, co-varying patterns of imaging signal that are combined, in varying proportions, to explain the observed data at each voxel.

In the neuroimaging context, each column of W describes how a single component loads across the m imaging modalities, capturing the multimodal signature of that component. Each row of H describes how that same component is spatially distributed across the n voxels of the brain. Together, the k components tile the multimodal signal space in a compact and interpretable way: rather than representing each voxel independently, NMF identifies a small number of recurrent, modality-spanning patterns whose weighted combination is sufficient to approximate the full input matrix.

In contrast to other potential integration methods (e.g., multiplying or summing up all single-modality analysis results), NMF identifies dominant patterns of covariance, enabling the discovery of latent disease factors rather than simple spatial convergence (10). Crucially, due to the non-negativity constrain, this approach avoids the problem of mutual cancellation that is inherent in simple summation or Principal Component Analysis (PCA) and therefore facilitates biological interpretation (7).

*Statistical evaluation of NMF-derived component maps*

Conventional parametric and non-parametric statistical tests can produce inflated significance estimates when comparing spatial brain maps because neighboring voxels are not statistically independent. This property, known as spatial autocorrelation, violates assumptions underlying most standard statistical frameworks (12,13). To address this issue, statistical approaches that preserve the spatial autocorrelation structure of the original data have been developed (12–14). Although these methods are typically used to assess the spatial correspondence between two brain maps (15), we adapted this framework to evaluate the statistical significance of voxel-wise contributions to the OPNMF-derived component maps (Fig. 1B).

For each of the seven single-modality meta-analytic *z*-maps, background voxels were excluded by setting them to NaN. The images were then resampled to 3 mm isotropic resolution using *fslmaths* to reduce computational demands of surrogate map computations. Next, Brain Surrogate Maps with Autocorrelated Spatial Heterogeneity (BrainSMASH; https://brainsmash.readthedocs.io/en/latest/index.html) was used to generate 1,000 surrogate maps for each modality (14,15). These surrogate maps preserve the spatial autocorrelation structure of the original brain map while randomizing the spatial arrangement of voxel values (47). To generate the surrogate maps, we first calculated the Euclidean distance matrix between all non-NaN cortical gray matter voxels within the SDM FreeSurfer mask, which was also resampled to 3 mm isotropic resolution. This distance matrix served as input for BrainSMASH to generate the surrogate maps. The fit between the spatial autocorrelation of the original and surrogate maps was assessed using variograms (14).

To establish voxel-wise null distributions for the OPNMF component outputs, OPNMF was performed once on the original dataset and separately on each of the 1,000 surrogate datasets, resulting in 1,001 OPNMF runs in total. Before decomposition, input maps were standardized across modalities using z-score normalization. To fulfill the non-negativity constraint of OPNMF, all values were subsequently shifted by subtracting the global minimum across the dataset, resulting in a minimum value of zero.

Statistical significance was assessed separately for every voxel in each OPNMF-derived component map. For each voxel, the observed weight from the original OPNMF solution was compared with the corresponding null distribution obtained from the 1,000 surrogate OPNMF runs. Empirical p-values were calculated as the proportion of surrogate voxel weights that exceeded the observed voxel weight. Voxels with empirical $p < 0.05$ were retained in the final component maps.

##### *Principal Component Analysis (PCA) as a control analysis for multi-modal integration*

To evaluate whether the multi-modal patterns identified by OPNMF were robust to the choice of dimensionality reduction method, we performed a control analysis using PCA (implemented in Python using *scikit-learn*, version 1.8.0).

Stacked meta-analysis z-maps for the seven investigated modalities were used as input for PCA after masking non-cortical voxels. Because PCA does not require non-negative input values, we tested two preprocessing strategies to enable comparability with the NMF framework. First, the input matrix was converted to positive values by taking the absolute value of all entries (“pca-abs”). Second, we applied the same preprocessing approach used for the NMF analyses: modality-wise *z*-score normalization

followed by subtraction of the global minimum value to shift all entries into the non-negative range while preserving the relative distribution of values across modalities ("pca-min").
PCA was then performed with two components. Component scores were projected back into volumetric space within SDM's FreeSurfer cortical mask and saved as NIfTI images for subsequent visualization and comparison with the OPNMF-derived spatial patterns.

*Comparison of multi-modal integration methods*

In order to compare the outputs of the different multi-modal integration methods (i.e., OPNMF, pca-abs, pca-min), all images were resampled to the same standard stereotactic space using *fslmaths* (i.e., MNI152NLIN6Asym, 2 mm resolution). Next, spatial Spearman correlation implemented in the *neuromaps* toolbox (https://netneurolab.github.io/neuromaps/index.html) was used to compare the overlap between two brain maps. BrainSMASH (n=1,000 permutations) (14,15) was used to assess statistical significance as described in the Supplementary Methods.

**Linking multi-modal macroscopic brain abnormalities with cell type expression**

*Microarray gene expression*

The Allen Human Brain Atlas (AHBA; https://human.brain-map.org/) provides a publicly available, high-resolution resource of regional gene expression in the human brain. For this atlas, bulk-tissue mRNA-based gene expression was determined across the whole brain from 6 postmortem human specimen (16). We used this unique data source as an estimate for cell-type specific gene expression across the brain. For this paper, the *abagen* toolbox (version 0.1.3; https://github.com/rmarkello/abagen; (17)) was used to correctly match AHBA gene expression data with native donor space MRI images following published recommendations (18) as follows:

- Probe aggregation: Microarray probes were aggregated to genes using data provided by Arnatkevičiutė et al. (18). Probes not matched to a valid Entrez ID were discarded. When multiple probes indexed the expression of the same gene, we selected the probe with the highest mean correlation across donor pairs (i.e., differential stability (19))
- Intensity-based filtering: Probes were filtered based on their expression intensity relative to background noise (20). Probes with intensity less than the background in at least 50 % of the samples across donors were discarded.
- Distance threshold: Samples were assigned to brain regions, using Montreal Neurological Institute (MNI) coordinates generated via nonlinear registrations (https://github.com/chrisfilo/alleninf), by minimizing the Euclidean distance between the MNI coordinates of each sample and the nearest surface vertex. Samples where the Euclidean distance to the nearest vertex was more than 2 standard deviations above the mean distance for all samples belonging to that donor were excluded. To reduce the potential for

misassignment, sample-to-region matching was constrained by hemisphere and gross structural divisions (i.e., cortex, subcortex/brainstem, and cerebellum, such that e.g., a sample in the left cortex could only be assigned to an atlas parcel in the left cortex) (18). All tissue samples not assigned to a brain region in the provided atlas were discarded.

- Sample normalization: To address inter-subject variation, tissue sample expression values were normalized across genes using a scaled robust sigmoid function (21).
- Gene normalization: Genes were normalized across tissue samples, again using a scaled robust sigmoid.
- Regional gene expression matrix: Samples assigned to the same brain region were averaged separately for each donor and then across donors, yielding a regional expression matrix in the 68-region surface-based Desikan-Killiany parcellation (22).

*Details on spin tests*

A null distribution of correlation coefficients between the two brain maps was computed by applying random rotations to spherical representations of one cortical surface map. Thereby, the intrinsic spatial autocorrelation structure of the brain map is preserved by assigning original values to the nearest rotated coordinates (12,13). The observed correlation coefficient was then compared against this null distribution to assess if spatial distributions of convergent abnormalities and cell type-specific gene expression were significantly linked.

**Linking multi-modal macroscopic brain abnormalities with evolutionarily cortices under recent evolutionary change**

*Map of a cortical representation of human-specific connectivity*

A cortical representation of human-specific connectivity was obtained from previous work by van den Heuvel et al. (23). In that study, diffusion-weighted imaging (DWI) and T1-weighted scans of 22 chimpanzees (*Pan troglodytes*, 29.4 ± 12.8 years) and age-matched 58 humans (*Homo sapiens*, 42.5 ± 9.8 years) (24) were analyzed to compare structural connectivity across species. The cortex was parcellated into a 114-region subdivision of the Desikan-Killiany atlas, also referred to as the Cammoun-2012 atlas (22,25). Deterministic fiber tracking was employed to identify connections between these regions, and fractional anisotropy was used as a proxy for connectivity strength of reconstructed connections. Human-specific connections were defined using a strict prevalence-based thresholding approach to minimize false positives. Specifically, connections had to be present in at least 60% of the human subject while being entirely absent (0% prevalence) in the chimpanzees. Using these criteria, the authors identified 27 human-specific connections, primarily involving the language network, orbitofrontal cortex, and parietal and temporal associative regions (23).

For the present study, we obtained a cortical map from the authors that represents the number of human-specific connections associated with this area for each region in the Cammoun-2012 atlas (25) (see Fig. 5A).

*Alternative map of cortices with human-specific connectivity*

To examine different definitions of human-specific connectivity, we also retrieved a different previously released map reflecting evolutionary modifications of brain connectivity (26). Instead of reflecting differences in discrete region-to-region white matter connectivity like the map used in the main analysis (23), Bryant and colleagues focused on identifying cortical areas where the overall “profile” of connectivity has diverged the most significantly from chimpanzees (Supplementary Figure S6) (26).

They analyzed *in vivo* diffusion-weighted imaging (DWI) and T1-weighted scans of 30 humans (aged 22 - 35 years) from the Human Connectome Project (27) and 23 chimpanzees (aged 26 ± 11 years). For each species and each point of the cortical surface, a vector of connectivity probabilities with 18 major white matter fiber bundles that are known to be homologous across species was reconstructed. By placing both species into a "common connectivity space," the authors quantified evolutionary divergence by calculating the Kullback–Leibler (KL) divergence between the connectivity profile of each human vertex and its best-matching vertex in the chimpanzees. This method resulted in a continuous spatial map of connectivity divergence, where higher values indicate "hotspots" of human specialization that are not represented in our closest relatives. These regions were primarily concentrated in the middle temporal lobe, the temporoparietal cortex, and the lateral frontal cortex (26).

For the present study, we obtained this vertex-wise divergence map (fsLR surface space with 32k density), which represents the degree to which regional connectivity architecture has been modified in the human lineage. To assess spatial correspondence between the connectivity divergence map and macroscopic brain aberrations related to schizophrenia, we first resampled the brain aberration map from MNI152 to fsLR space using neuromaps to match the connectivity divergence map. Spin tests were used for statistical evaluation of the spatial Spearman correlation between the two maps (12,15).

*Spatial overlap analysis between the cortical representation of human-specific connectivity and multi-modal brain abnormalities*

To examine the spatial correlation between the cortical representation of human-specific connectivity and the convergent multi-modal macroscopic brain abnormalities in schizophrenia, we first parcellated the W component maps from OPNMF thresholded for significance into 114 cortical regions using the Cammoun-2012 atlas (25) to match the representation of the map representing human-specific connections ending in the respective cortical region. Next, the ROI-wise Dice coefficient between the two binarized maps was computed to assess the spatial overlap. To assess statistical significance, we performed a permutation test (n = 10,000 permutations), in which the number of regional values of human-specific connectivity endpoints were randomly reassigned across cortical regions in each iteration. By recalculating the Dice coefficient of the permutated map with macroscopic brain aberrations related to schizophrenia, a null distribution of Dice coefficients was determined. The empirical p-value was determined by comparing the observed Dice coefficient against the null distribution generated with the permutation (Fig. 5A). As a control analysis for investigating the association between the cortical representation of human-specific connectivity and multi-modal brain abnormalities, in addition to the Dice score, we also calculated spatial Spearman correlations. Spatially constrained null models for volumetric data, i.e., BrainSMASH, n=1,000 permutations, permutation of the integrated meta-analysis map) were used to assess statistical significance of the results (14,15).

*Map of human-specific cortical expansion*

Cortical representations of human-specific cortical expansion were obtained from Wei and colleagues (28) (Fig. 5B). In that study, T1-weighted images from 29 adult chimpanzees and 50 adult human subjects derived from the Human Connectome Project (https://www.humanconnectome.org) were processed using FreeSurfer (v5.3.0; https://surfer.nmr.mgh.harvard.edu/) for tissue classification and cortical surface reconstruction (22). Vertex-to-vertex mapping across chimpanzee and humans and subsequent computation of vertex-wise and region-wise expansion was performed on pial surface reconstructions (28). For the present study, we obtained a cortical map from the authors that was in fsaverage space with 164k density. To reduce computational load, the map was downsampled to 10k density. So was the meta-analysis map.

*Spatial overlap analysis between human-specific cortical expansion and multi-modal brain abnormalities*

To compare the spatial overlap between multi-modal macroscopic brain abnormalities of schizophrenia as derived with NMF and cortical expansion from humans to chimpanzees, both maps were resampled from MNI152 3 mm stereotactic space or from fsaverage5, respectively, to fsLR space with 32k density using *neuromaps* (15). The two resampled brain maps were compared using spatial Spearman correlation, while statistical significance was assessed using spatially constrained null models for surface-based data, i.e., “spin tests” with 1,000 permutations (12,15).

#### Supplementary Results

##### Description of the sample

Systematic literature research and study selection using PRISMA criteria (32,33) resulted in the inclusion of 209 studies in total. Supplementary Figures S1 represent flowcharts of the screening process for each investigated modality. For ALFF, 28 studies comprising 2287 patients with schizophrenia (mean age: 27.24 years, mean illness duration: 40.14 months, mean PANSS-positive: 19.60, mean PANSS-negative: 19.80, mean PANSS-general: 41.82, mean PANSS-total: 82.14) and 2107 healthy controls (mean age: 27.05 years) were identified. For ReHo, 25 publications met the inclusion criteria, comprising 1211 patients (mean age: 27.61 years, mean illness duration: 55.34 months, mean PANSS-positive: 19.88, mean PANSS-negative: 20.78, mean PANSS-general: 40.28, mean PANSS-total: 82.02) and 1057 HC (mean age: 28.54 years). Fourteen studies examining rCBF were included, encompassing 541 patients (mean age: 34.00 years, mean illness duration: 118.94 months, mean PANSS-positive: 17.78, mean PANSS-negative: 17.69, mean PANSS-general: 34.23, mean PANSS-total: 71.20) and 452 controls (mean age: 33.35 years). Four experiments investigated FDG-PET, with a total of 111 patients (mean age: 30.23 years, mean illness duration: 62.48 months, mean PANSS-positive: 18.45, mean PANSS-negative: 19.60, mean PANSS-general: 37.50, mean PANSS-total: 75.55) and 70 controls (mean age: 29.73 years). For CTh, 21 articles met the inclusion criteria, which investigated a total of 2568 patients (mean age: 31.12 years, mean illness duration: 136.56 months, mean PANSS-positive: 16.32, mean PANSS-negative: 17.66, mean PANSS-general: 33.97, mean PANSS-total: 70.50) and 1633 HC (mean age: 29.01 years). Eleven publications examined gyrification between a total of 868 patients (mean age: 29.29 years, mean illness duration: 82.68 months, mean PANSS-positive: 15.69, mean PANSS-negative: 17.07, mean PANSS-general: 28.91, mean PANSS-total: 65.55) and 695 controls (mean age: 28.95 years). For VBM with modulation, 106 studies were included in the present meta-analysis, comprising 4885 patients (mean age: 32.14 years, mean illness duration: 105.66 months, mean PANSS-positive: 18.45, mean PANSS-negative: 19.59, mean PANSS-general: 37.59, mean PANSS-total: 74.87) and 4863 HC (mean age: 31.21 years).

##### Heterogeneity and potential publication bias

To assess the robustness of the results, we performed analyses of heterogeneity and potential publication bias (Supplementary Tables S3). Across modalities, most peaks did not show significant heterogeneity (i.e., $I^2$-statistic << 50) or potential publication bias (i.e., Egger's test p-value >> 0.05). Only one positive peak for ALFF (MNI coordinates (18,12,10), $I^2$-statistic = 71.20) exhibited a $I^2$-statistic of more than 50%, indicating high between-study heterogeneity. Furthermore, one negative peak of rCBF (MNI coordinates (36,-90,6), Egger's test p-value < 0.001) as well as one negative peak of VBM (MNI coordinates (40,-50,-22), Egger's test p-value < 0.001) showed a significant p-value in the Egger's test, suggesting significant asymmetry in the funnel plots, which can be interpreted as potential publication bias. The Egger's test for potential publication bias for the meta-analysis of FDG-PET studies could not be successfully conducted due to the small number of included studies (n = 4). Overall, these findings suggest overall high homogeneity of studies that is likely not influenced by publication bias.

**Control analyses for multi-modal integration**

*OPNMF using K = 3*

To assess the robustness of the OPNMF solution against the choice of the number of components, we repeated the multimodal convergence analysis using K = 3. The resulting component maps are depicted in Supplementary Figure S4. The convergent component from the K = 2 (i.e., component 2) solution was nearly identical in its spatial topography to the K = 3 solution (Spearman's $\rho = 0.979$, $p_{SMASH} = 0.001$), demonstrating the stability of the identified insular and mediofrontal convergence pattern across parameters.

*Substituting OPNMF with PCA*

To further evaluate the robustness of the multimodal integration approach, we substituted OPNMF with PCA. As detailed in the Supplementary Methods, two input matrix preprocessing strategies were compared: using the absolute value of all entries (PCA-abs) or modality-wise z-score normalization followed by subtraction of the global minimum value (PCA-min), both ensuring comparability with the OPNMF framework by enforcing non-negativity. Spatial correspondence between component 2 of the K = 2 OPNMF solution and the first principal component was moderate for PCA-abs (Spearman's $\rho = 0.735$, $p_{SMASH} = 0.001$, Supplementary Figure S5) and very high for PCA-min (Spearman's $\rho = -0.950$, $p_{SMASH} = 0.001$) with the negative sign reflecting a sign flip inherent to PCA rather than a substantive difference in spatial topography. Collectively, these results indicate high stability of the convergence of schizophrenia-related abnormalities in mediofrontal and insular regions, independent of the multimodal integration method selected.

#### Supplementary Figures

**Figures S1: Flowcharts of study selection**

*Figure S1a: PRISMA flowchart study selection for ALFF*

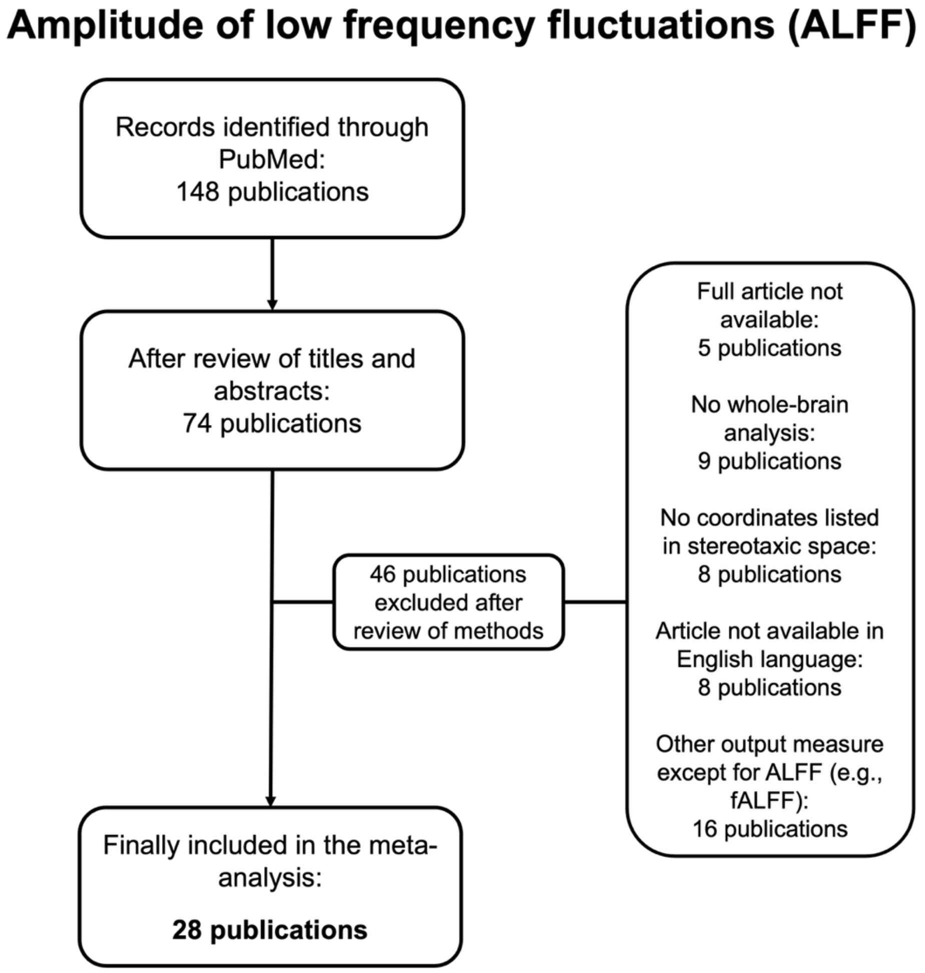

*Figure S1b: PRISMA flowchart study selection for ReHo*

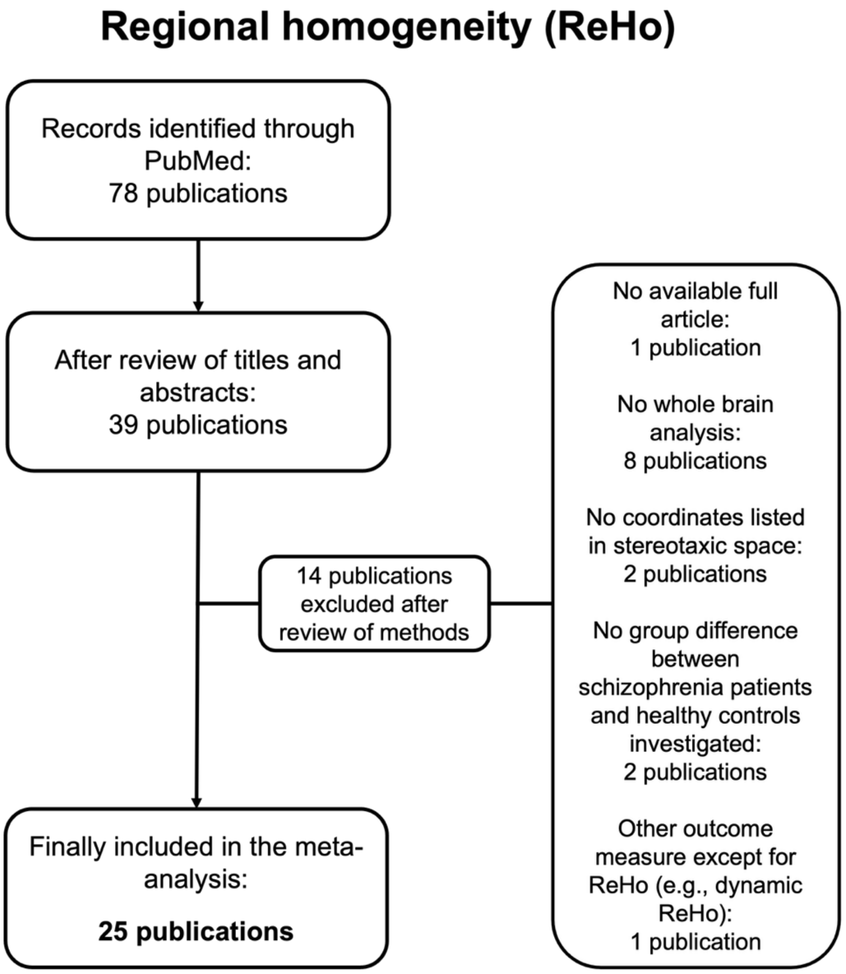

*Figure S1c: PRISMA flowchart study selection for rCBF*

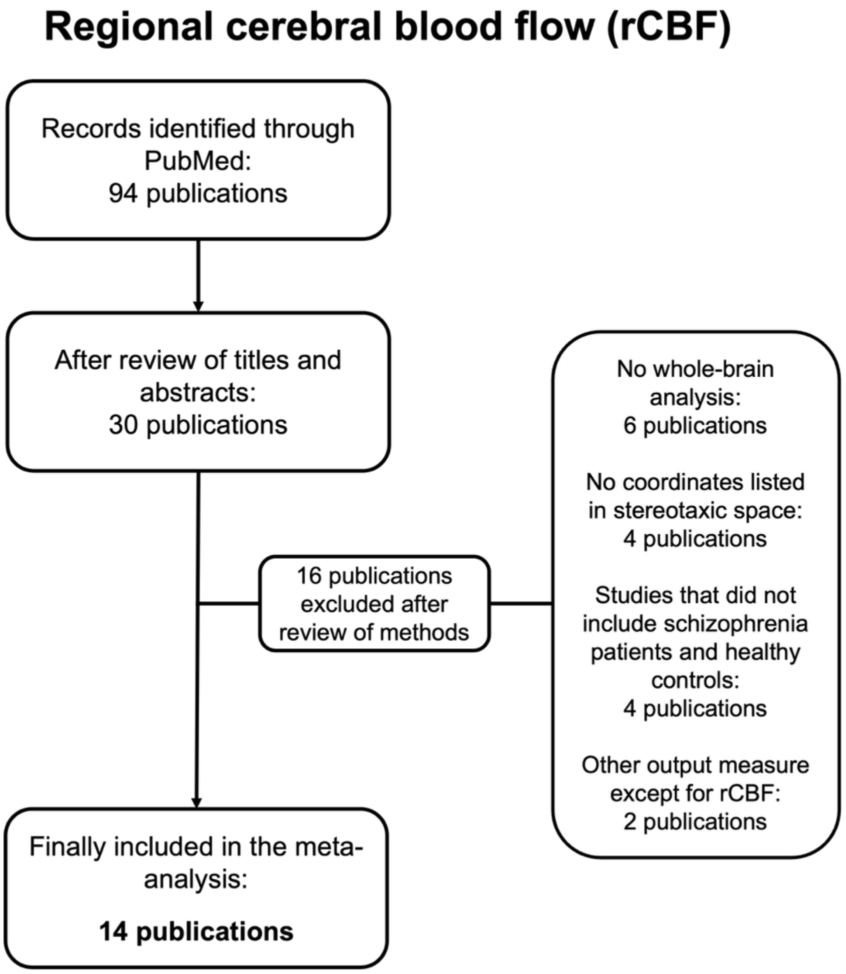

*Figure S1d: PRISMA flowchart study selection for glucose metabolism*

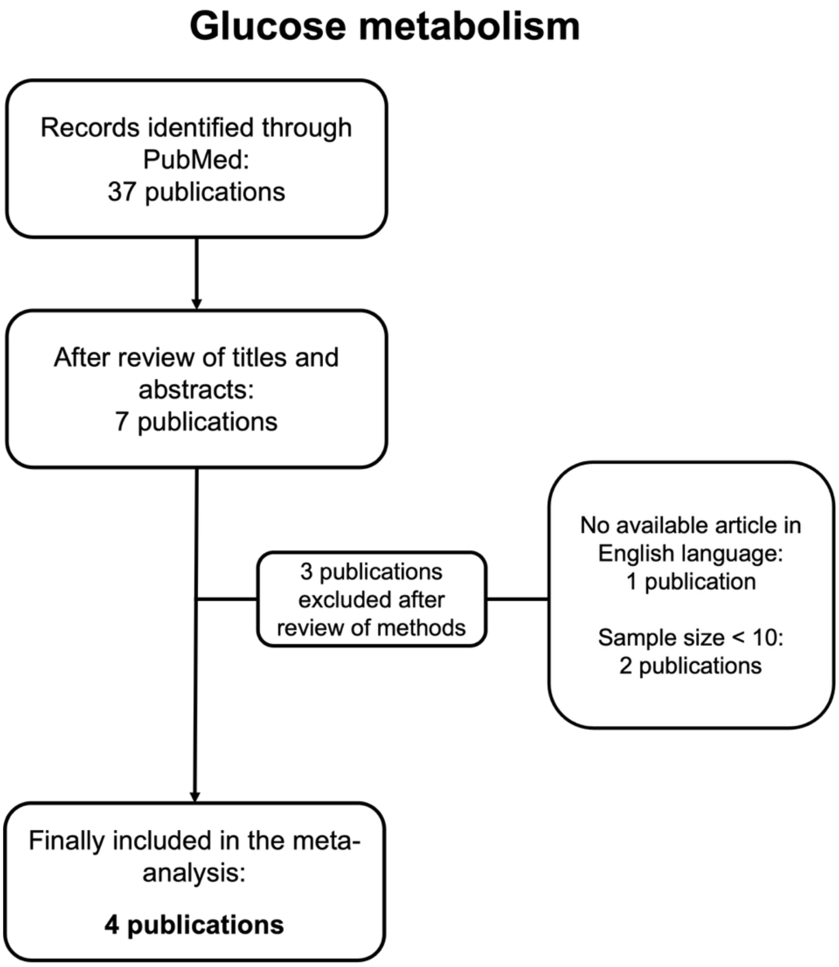

*Figure S1e: PRISMA flowchart study selection for CTh*

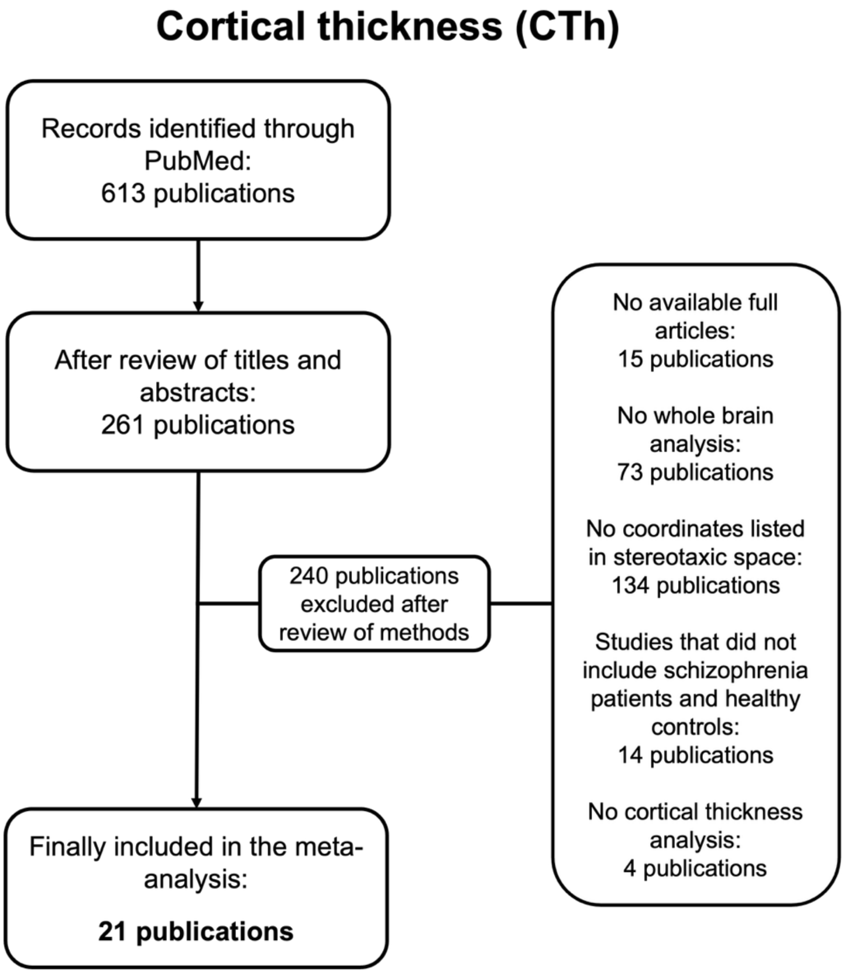

*Figure S1f: PRISMA flowchart study selection for gyrification*

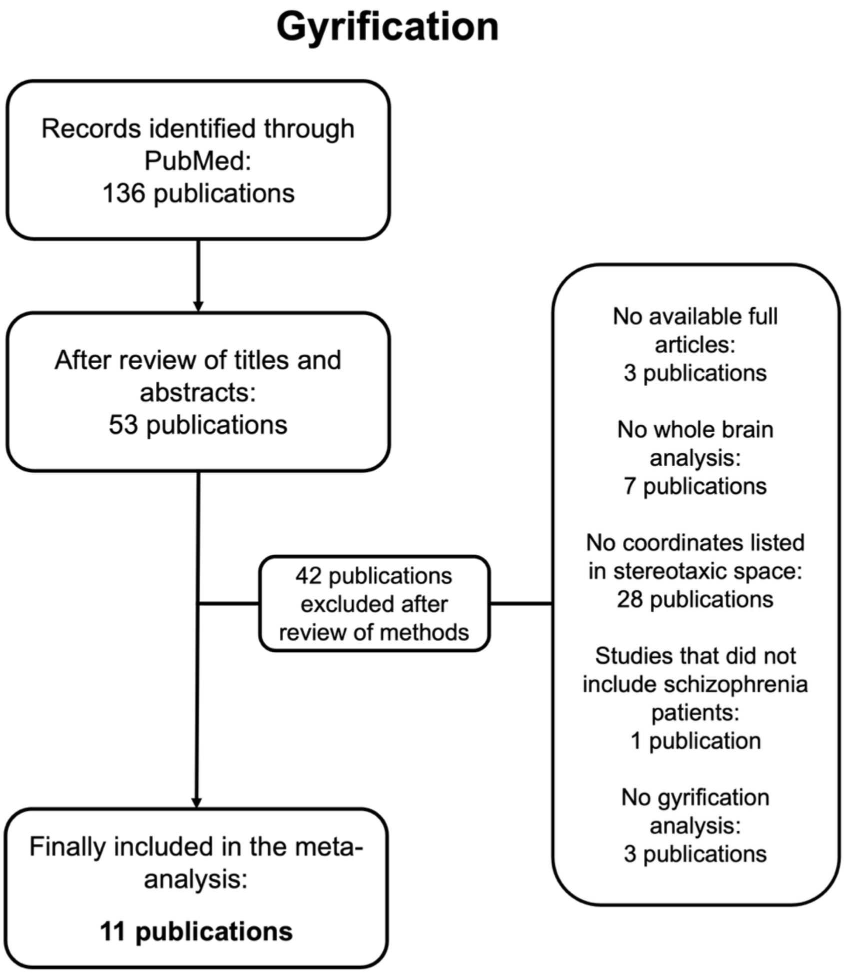

*Figure S1g: PRISMA flowchart study selection for GMV*

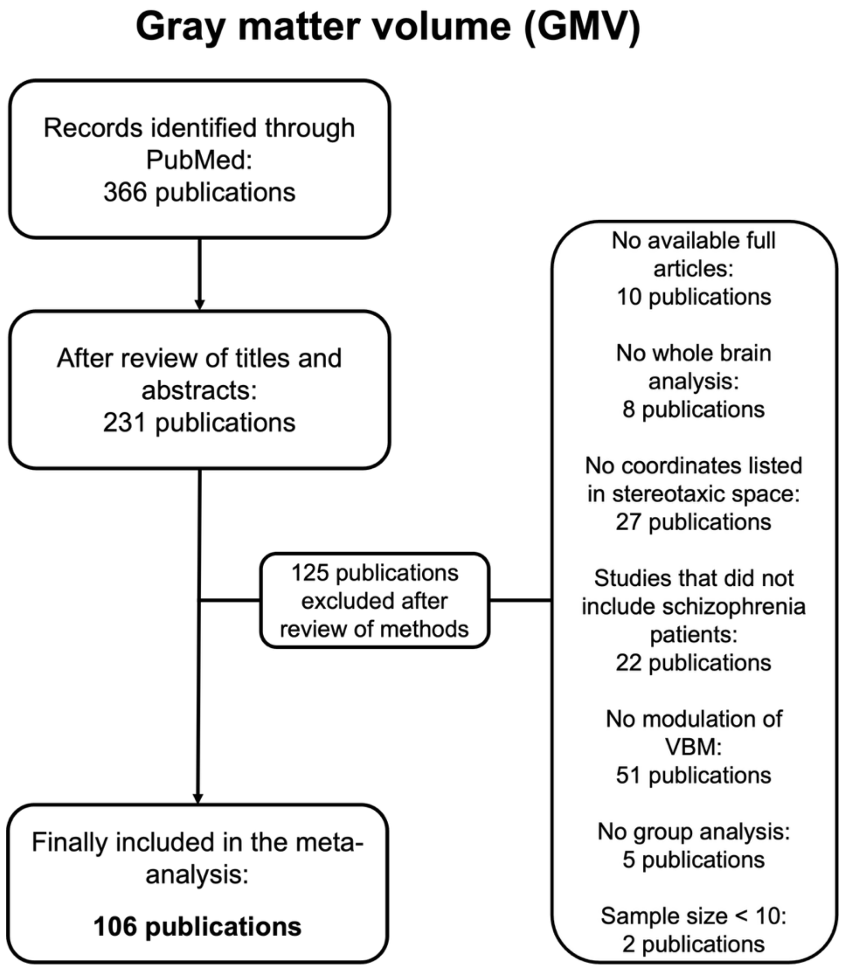

**Supplementary Figure S2: Unthresholded outcome maps of the seven meta-analyses for each modality**

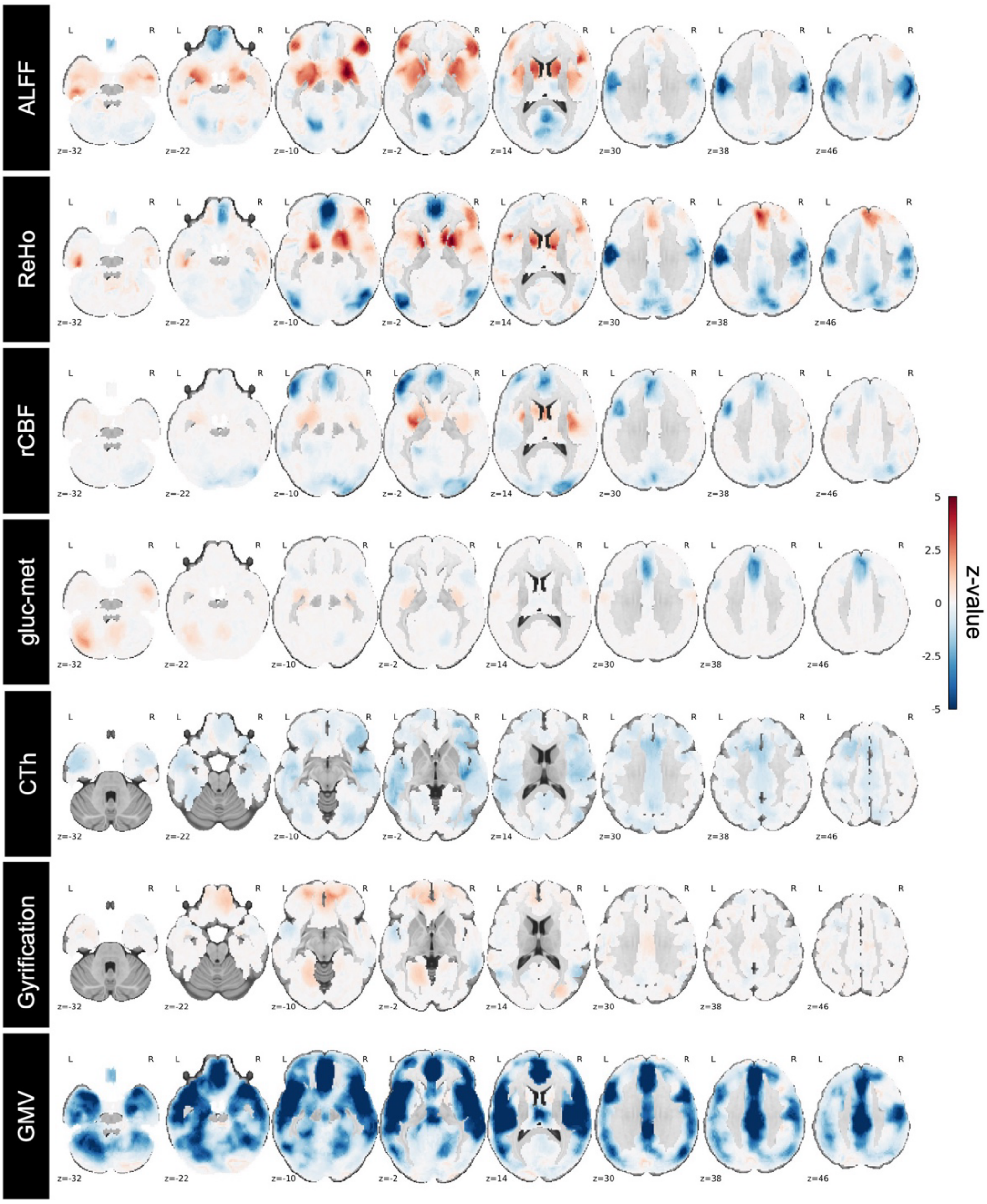

*Supplementary Figure S2: Outcome maps of the seven meta-analyses for each modality, representing the difference between patients with schizophrenia and healthy controls presented as z-values. Investigated modalities included amplitude of low-frequency fluctuations (ALFF), regional homogeneity (ReHo), regional cerebral blood flow (rCBF), glucose metabolism (gluc-met) based on [$^{18}$F]-Fluorodeoxyglucose positron emission tomography, cortical thickness (CTh), gyrification, and gray matter volume (GMV) based on voxel-based morphometry with modulation. Results are presented as unthresholded z-values. Maps thresholded for significance (p<0.001) are visualized in Figure 2.*

**Supplementary Figure S3: Schematic representation of non-negative matrix factorization (NMF)**

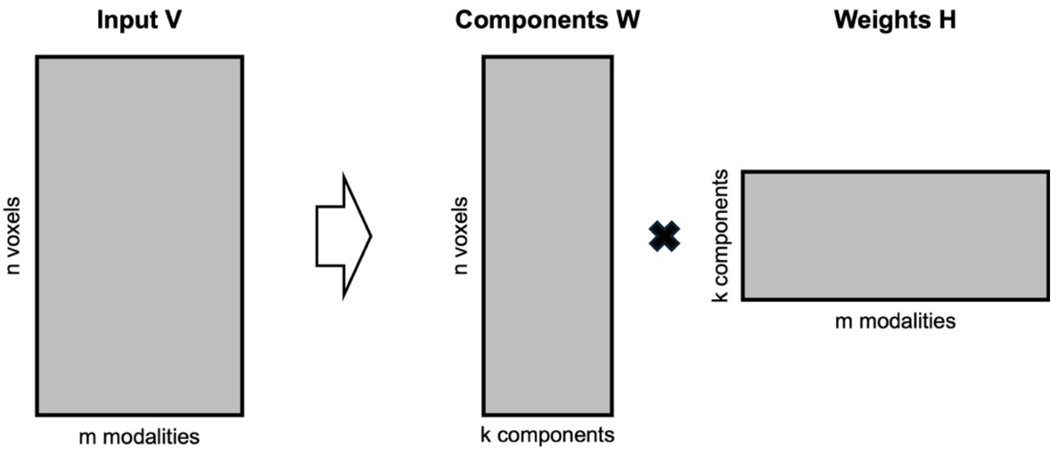

*Supplementary Figure S3: Schematic representation of non-negative matrix factorization (NMF). NMF decomposes an input matrix V into a component matrix W and a weight matrix H, so that the reconstruction error between input V and reconstructed input W x H is minimized (7).*

**Supplementary Figure S4: Component maps for NMF with K = 3**

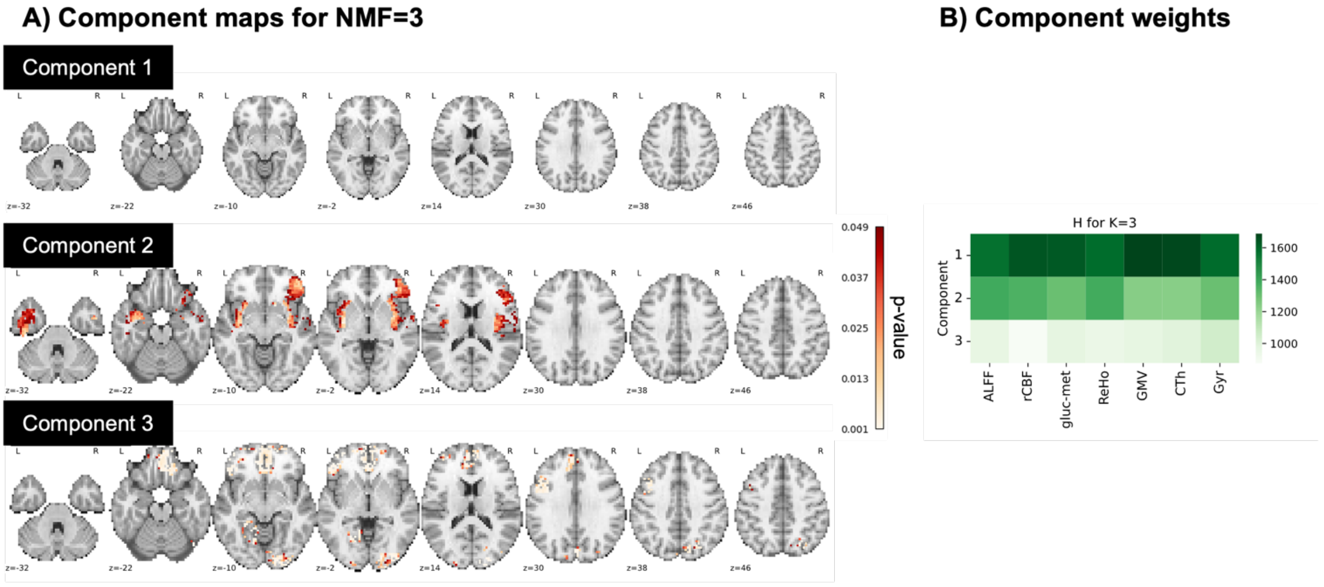

*Supplementary Figure S4: Component maps for NMF with K=3, thresholded with $p_{SMASH}$<0.05.* ***(A)*** *The spatial maps of resulting components are visualized. Component map 1 did not include any significant voxels.* ***(B)*** *The component weights for each imaging modality are presented. Amplitude of low-frequency fluctuations (ALFF), regional cerebral blood flow (rCBF), glucose metabolism (gluc-met) based on [$^{18}$F]-Fluorodeoxyglucose positron emission tomography (FDG-PET), regional homogeneity (ReHo), gray matter volume (GMV) based on voxel-based morphometry with modulation, cortical thickness (CTh), and gyrification (Gyr).*

#### Supplementary Figure S5: Multimodal integration using principal component analysis

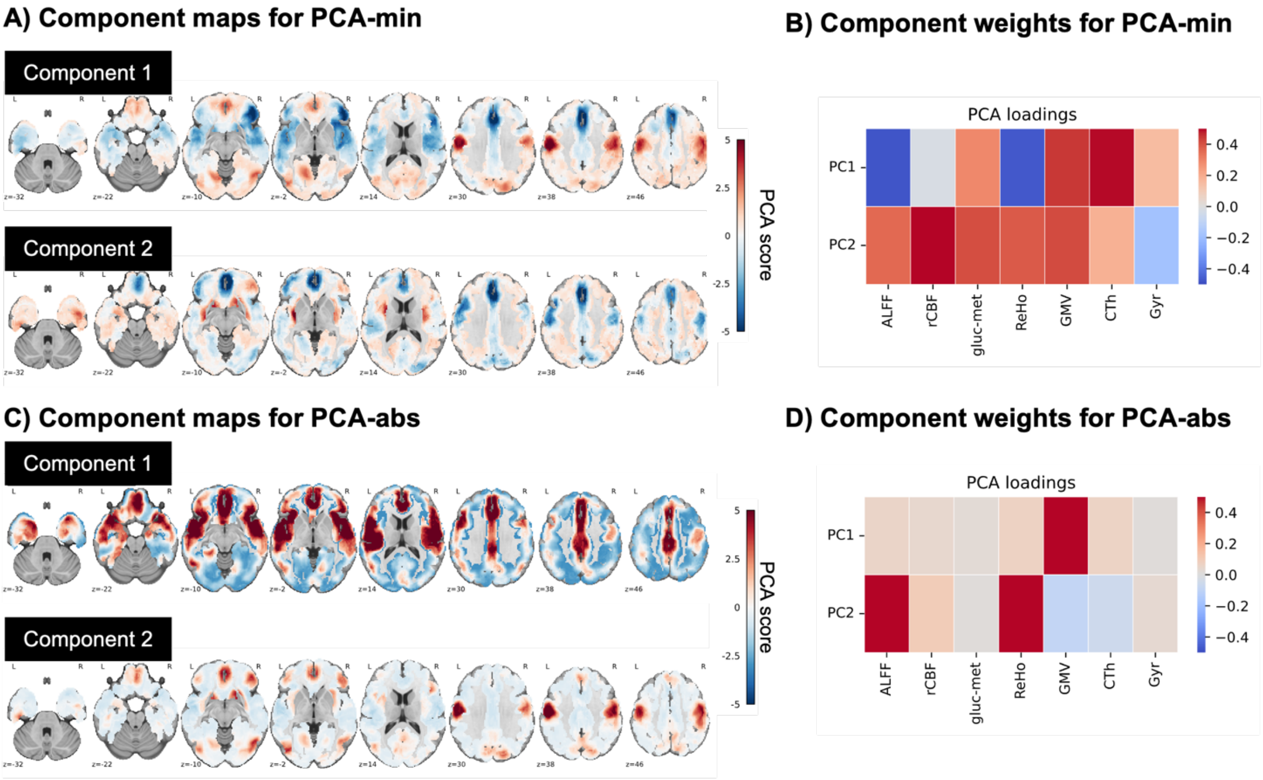

*Supplementary Figure S5: Multimodal integration using principal component analysis (PCA). On the left, component maps of the first two principal components are visualized on MNI152 templates. Color-coding represents PCA scores. On the right, PCA loadings are depicted. Two different preprocessing methods were compared: A) PCA-min, B) PCA-abs.*

**Supplementary Figure S6: Cortical representation of human-specific connectivity profile divergence**

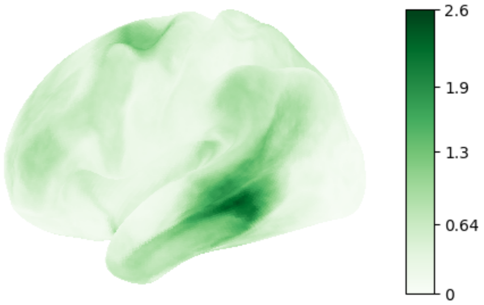

*Supplementary Figure S6: Cortical representation of human-specific connectivity profile divergence between humans and chimpanzees by Bryant and colleagues (26). Data are projected on cortical surfaces in fs-LR space with 32k density. Compared to the cortical representation of human-specific connectivity used in the main analysis (23), the alternative map depicted here focuses on the profile of connectivity of a given vertex with each of the white matter tracts (26).*

### Supplementary Tables

##### Tables S1: Demographic and clinical information of included studies

*Table S1a: Demographic and clinical information of included studies for ALFF*

| Study | Control subjects | | Schizophrenia subjects | | | | | | |
|---|---|---|---|---|---|---|---|---|---|
| | n | Mean age [years] | n | Mean age [years] | Illness duration [months] | PANSS positive | PANSS negative | PANSS general | PANSS total |
| Alonso-Solis, 2017 (31) | 20 | 37.75 | 19 (hallucinatory) | 40.05 | 193.32 | 17.89 | 21.47 | 34.22 | |
| | | | 14 (non-hallucinatory) | 36.43 | 96 | 11.43 | 14.36 | 27.36 | |
| Chen, 2022 (32) | 235 | 28.17 | 89 (medicated) | 27.65 | 43.07 | | | | |
| | | | 81 (unmedicated) | 25.53 | 8.84 | | | | |
| Chen, 2024 (33) | 73 (adults) | 22.4 | 84 (adults) | 22.7 | 12.4 | 20.6 | 22.8 | 44.1 | 87.5 |
| | 46 (adolescents) | 15.2 | 66 (adolescents) | 15.5 | 12.1 | 19.7 | 22 | 41.2 | 83 |
| Chen, 2025 (34) | 30 | 38.67 | 40 (combined treatment) | 38.98 | 12.5 | 9 | 10.5 | 24 | 42.5 |
| | | | 28 (risperidone-treated) | 38.39 | 11 | 8 | 11.5 | 26.5 | 50 |
| Cui, 2016 (35) | 19 | 23.79 | 17 (AVH) | 21.24 | 6.51 | 31.12 | 25.53 | 49.59 | 106.24 |
| | | | 19 (non-AVH) | 22.53 | 10.2 | 17.93 | 22.73 | 47.4 | 88.07 |
| Gao, 2022 (36) | 128 | 30.33 | 131 (medicated) | 26.34 | 83.04 | | | | 89.63 |
| | 50 | 23.48 | 49 (unmedicated) | 22.69 | 22.45 | 22.27 | 22.82 | 46.22 | 91.31 |
| Hare, 2017 (37) | 155 | 37.8 | 42 (AH) | 37.8 | 18 | 16.6 | 13.7 | | 57.7 |
| | | | 40 (AVH) | 37.2 | 17 | 17.6 | 15.2 | | 63.3 |
| | | | 61 (non-hallucinatory) | 40.2 | 17.3 | 12.9 | 13.9 | | 54 |

| | | | | | | | | | |
|---|---|---|---|---|---|---|---|---|---|
| Huang, 2010 (38) | 66 | 24.5 | 66 | 24.2 | 8.8 | 26.4 | 20.7 | 51.3 | 107.2 |
| Kuang, 2022 (39) | 33 | 24.2 | 28 | 25.1 | 8.7 | 17.4 | 13.6 | | 65.1 |
| Lei, 2015 (40) | 50 (males) | 24.8 | 61 (males) | 24.31 | 8.19 | 14.74 | 16.58 | 56.27 | 87.59 |
| | 52 (females) | 24.71 | 63 (females) | 24.62 | 5.5 | 15.77 | 14.85 | 58.65 | 89.27 |
| Li, 2016 (41) | 16 | 22.4 | 20 | 22.9 | 6.4 | 25.1 | 18.8 | 49.7 | 101.6 |
| Li, 2017 (42) | 42 | 23.29 | 41 (deficit) | 23.32 | 20.15 | 22.15 | 27.49 | 46.63 | 99.38 |
| | | | 42 (non-deficit) | 22.86 | 19.83 | 25.38 | 16.48 | 43.36 | 86.22 |
| Li, 2023 (43) | 81 | 30.93 | 88 | 28.76 | 74.84 | 13.22 | 25.75 | | 93.16 |
| Liang, 2019 (44) | 30 | 12.9 | 30 | 13 | 5.5 | | | | 74.5 |
| | | | 21 (with OC symptoms) | 13.2 | 5.4 | | | | 76.4 |
| Liang, 2024 (45) | 33 | 14.33 | 50 | 14.22 | 5.26 | | | | |
| Liu, 2016 (46) | 27 | 27.44 | 27 | 25.44 | 18.32 | 21.56 | 23.15 | 41.3 | 85.78 |
| Liu, 2021 (47) | 153 | 26.59 | 153 | 27.05 | 41.19 | 24.66 | 19.37 | 46.18 | 90.21 |
| Lui, 2010 (48) | 34 | 25 | 34 | 24.6 | 7.8 | 26.9 | 19.1 | 49.9 | 104.2 |
| Lui, 2015 (49) | 59 | 38 | 37 | 36 | 177.72 | 18.2 | 18.31 | 34.69 | 71.2 |
| Ren, 2013 (50) | 100 | 24.39 | 100 | 24.3 | 6.25 | 25.11 | 18.84 | 47.63 | 97.88 |
| Salvador, 2017 (51) | 122 | 36.51 | 116 | 36.76 | 178.56 | 16.57 | 19.57 | 33.08 | 69.23 |
| Shao, 2022 (52) | 18 (Center 1) | 24 | 9 (Center 1, severely impaired) | 22.89 | 19.11 | 18.44 | 29.44 | 41.78 | 89.67 |
| | | | 25 (Center 1, moderately impaired) | 24.8 | 10.08 | 16.84 | 22.32 | 42.72 | 81.88 |

| | | | | | | | | | |
|---|---|---|---|---|---|---|---|---|---|
| | 36 (Center 2) | 28.58 | 7 (Center 2, severely impaired) | 28.57 | 21.72 | 25 | 28.57 | 47.43 | 101 |
| | | | 31 (Center 2, moderately impaired) | 29.52 | 16.26 | 26.45 | 21.23 | 44.61 | 92.26 |
| | 43 (Center 3) | 22.49 | 47 (Center 3, severely impaired) | 23.32 | 3.95 | 24.09 | 26.38 | 47.4 | 97.87 |
| | | | 36 (Center 3, moderately impaired) | 22.22 | 4.78 | 21.5 | 23.08 | 44.11 | 89.23 |
| Turner, 2013 (53) | 176 | 37.5 | 186 | 38.9 | 157.2 | 15.9 | 15.4 | 28.8 | 60.2 |
| Xie, 2021 (54) | 33 | 32.03 | 30 | 30.3 | 21.36 | 19.65 | 19.85 | 40.35 | 79.85 |
| Yang, 2020 (55) | 25 | 44.36 | 25 | 47.12 | 217.44 | 24.29 | 24.04 | 45.04 | 93.36 |
| Yin, 2021 (56) | 30 | 20.8 | 30 | 19.8 | | 21.8 | 18.8 | 37.7 | 78.3 |
| Yu, 2014 (57) | 62 | 29.9 | 69 | 31.7 | 85.2 | 12.1 | 13.4 | 27.4 | 52.9 |
| Zheng, 2016 (58) | 30 | 15.43 | 35 | 15.5 | 6.6 | 20.42 | 20.91 | 33.28 | 74.62 |

*Note*: AH auditory hallucinations; AVH auditory-verbal hallucinations; OC obsessive-compulsive

*Table S1b: Demographic and clinical information of included studies for ReHo*

| Study | Control subjects | | Schizophrenia subjects | | | | | | |
|---|---|---|---|---|---|---|---|---|---|
| | n | Mean age [years] | n | Mean age [years] | Illness duration [months] | PANSS positive | PANSS negative | PANSS general | PANSS total |
| Bai, 2016 (59) | 17 | 28.71 | 17 | 26 | 40.29 | 15.29 | 24.88 | | 82.06 |
| Chen, 2013 (60) | 44 | 35.68 | 36 | 28.28 | | | | | 76.57 |
| Chen. 2025 (61) | 28 | 39.29 | 20 (metabolic syndrome) | 41.45 | 108 | | | | |
| | | | 23 (without metabolic syndrome) | 34.91 | 152.88 | | | | |
| Cui, 2016 (35) | 19 | 23.79 | 17 (AVH) | 21.24 | 6.51 | 31.12 | 25.53 | 49.59 | 106.24 |
| | | | 15 (non-AVH) | 22.53 | 10.2 | 17.93 | 22.73 | 47.4 | 88.07 |
| Fang, 2021 (62) | 34 | 21.35 | 34 (non-depressive) | 22.68 | 13.59 | 23.35 | 21.82 | 43.68 | 88.85 |
| | | | 35 (depressive) | 22.26 | 10.74 | 22.8 | 19.77 | 46.71 | 89.29 |
| Fang, 2024 (63) | 41 | | 33 | | | | | | |
| Gao, 2015 (64) | 14 | 34.9 | 14 | 33.2 | 110.4 | 16.4 | 22.6 | 30.8 | 74.1 |
| Gao, 2018 (65) | 29 | 32.73 | 17 (treatment resistant) | 31.24 | 168 | 27.53 | 21.05 | 49.12 | 97.76 |
| | | | 17 (non-treatment resistant) | 36.82 | 94.56 | 9.53 | 8.41 | 19.24 | 37.29 |
| Gao, 2020 (66) | 50 | 28.38 | 57 | 31.63 | 30.24 | 26.39 | 20.68 | 44.79 | 91.84 |
| Gou, 2018 (67) | 21 | 28.8 | 28 | 23.9 | 15.1 | 17.8 | 21 | 38.9 | 85.7 |

| | | | | | | | | | |
|---|---|---|---|---|---|---|---|---|---|
| Jin, 2021 (68) | 24 | 30.92 | 23 | 31.74 | | | | | |
| Jing, 2023 (69) | 51 | 41 | 56 | 41.76 | | 16.04 | 22.64 | 33 | 71.67 |
| Li, 2023 (43) | 81 | 30.93 | 88 | 28.76 | 74.84 | 13.22 | 25.75 | | 93.16 |
| Liu, 2006 (70) | 18 | 24.44 | 18 | 23.67 | 26.83 | | | | 80.39 |
| Liu, 2016 (46) | 27 | 27.44 | 27 | 25.44 | 18.32 | 21.56 | 23.15 | 41.3 | 85.78 |
| Liu, 2025 (71) | 56 | 27.71 | 56 | 28.29 | | | | | |
| Shan, 2021 (72) | 20 | 25.7 | 19 (drug plus psychotherapy) | 26.05 | | | | | |
| | | | 20 (drug therapy) | 22.75 | | | | | |
| Sun, 2021 (73) | 42 | 29.17 | 40 | 28.15 | 7.86 | | | | |
| Wang, 2018 (74) | 31 | 15.42 | 48 | 15.79 | 5.35 | 21.5 | 17.92 | 34.25 | 75.1 |
| Yan, 2020 (75) | 74 | 26.27 | 69 | 24.22 | 13.74 | 24.42 | 17.58 | 42.19 | 84.19 |
| Yang, 2021 (76) | 39 | 40.94 | 37 | 39.7 | 204 | 13.6 | 23.9 | | 76 |
| Yang, 2022 (77) | 41 (adolescents) | 15.57 | 49 (adolescent first episode) | 15.16 | | 23.36 | 22.9 | 38.05 | 87.15 |
| | 138 (adults) | 31.04 | 57 (adult first episode) | 25.93 | | 22.98 | 18.54 | 39.59 | 81.48 |
| | | | 51 (chronic) | 27.16 | | 23.44 | 19.41 | 39.67 | 82.68 |

| | | | | | | | | | |
|---|---|---|---|---|---|---|---|---|---|
| Yin, 2021 (56) | 30 | 20.8 | 30 | 19.8 | 8.5 | 21.8 | 18.8 | 37.7 | 78.3 |
| Yu, 2013 (74) | 62 | 29.9 | 69 | 31.7 | 85.2 | 12.1 | 13.4 | 27.4 | 52.9 |
| Zhao, 2019 (79) | 26 | 22.6 | 44 (deficit) | 23.7 | 12 | 15.3 | 24.7 | 62 | 102 |
| | | | 47 (non-deficit) | | | | | | |

*Note*: AVH auditory-verbal hallucinations

*Table S1c: Demographic and clinical information of included studies for rCBF*

| Study | Control subjects | | Schizophrenia subjects | | | | | | |
|---|---|---|---|---|---|---|---|---|---|
| | n | Mean age [years] | n | Mean age [years] | Illness duration [months] | PANSS positive | PANSS negative | PANSS general | PANSS total |
| Chen, 2022 (80) | 48 | 21.9 | 46 (AVH) | 21.7 | | 20.1 | 20.2 | 41.6 | 82.3 |
| | | | 39 (non-AVH) | 20.2 | | 18.1 | 21 | 42.6 | 81.6 |
| Cui, 2017 (80) | 25 | 26 | 25 (non-AH) | 24 | 18 | 21 | 23 | 48 | 92 |
| | | | 25 (AH) | 24 | 14 | 31 | 27 | 51 | 109 |
| Foucher, 2018 (82) | 27 | 39 | 20 (periodic catatonia) | 38 | 187.2 | 11.7 | 19.6 | 34.4 | 65.7 |
| | | | 9 (cataphasia) | 38 | 172.8 | 16.8 | 16.8 | 33.1 | 66.7 |
| Horn, 2009 (84) | 13 | 26.6 | 13 | 29.6 | 33.48 | 13.48 | 15.5 | 34.92 | 63.9 |
| Kindler, 2015 (86) | 27 | 39.8 | 34 | 41.5 | | 19.9 | 18.9 | 36.7 | 75.5 |
| Kindler, 2018 (88) | 31 | 39.4 | 32 | 41.6 | | 19.6 | 19.7 | 37.3 | 76.6 |
| Ota, 2014 (89) | 42 | 37.9 | 36 | 39.8 | 201.6 | 15.3 | 4.9 | 31.7 | 61.8 |
| Pinkham, 2011 (90) | 24 | 35.73 | 30 | 36.73 | | | | | |
| Pinkham, 2015 (91) | 25 | 33.64 | 16 (non-paranoid) | 38.81 | | 12.63 | 11.56 | 25.19 | 49.38 |
| | | | 16 (paranoid) | 38.56 | | 19.19 | 3.25 | 32 | |
| Scheef, 2010 (92) | 25 | 30 | 11 | 32 | | 20.2 | 21.4 | 1.5 | 43.1 |
| Sun, 2024 (93) | 16 | n.a. | 36 | 39.47 | 171.72 | 18.43 | 19.23 | 35.33 | 72.97 |
| Walther, 2011 (91) | 14 | 31.71 | 11 | 35.36 | 106.8 | 11.73 | 18 | 24.54 | 54.27 |

| | | | | | | | | | |
|---|---|---|---|---|---|---|---|---|---|
| Walther, 2017 (95) | 41 | 38.6 | 15 (with catatonia) | 35.9 | 153.4 | 16 | 21.9 | 39.2 | 77.1 |
| | | | 27 (without catatonia) | 37.1 | 126.4 | 17.9 | 16.4 | 32.8 | 67.1 |
| Zhu, 2015 (96) | 94 | 33.3 | 100 | 33.6 | 122.9 | 17 | 20.1 | 34.2 | 71.3 |

*Note*: AH auditory hallucinations; AVH auditory-verbal hallucinations

*Table S1d: Demographic and clinical information of included studies for FDG-PET*

| **Study** | **Control subjects** | | **Schizophrenia subjects** | | | | | | |
|---|---|---|---|---|---|---|---|---|---|
| | **n** | **Mean age [years]** | **n** | **Mean age [years]** | **Illness duration [months]** | **PANSS positive** | **PANSS negative** | **PANSS general** | **PANSS total** |
| Desco, 2002 (94) | 18 | 29.63 | 17 (FES) | 23.68 | 13.92 | | | | |
| | | | 34 (chronic) | 36.05 | 135.12 | | | | |
| Kim, 2017 (98) | 18 | 30.7 | 19 | 32.6 | 27.6 | 21 | 22.5 | 41.2 | 84.7 |
| Lehrer, 2005 (99) | 13 | 28.5 | 12 | 29 | 45.75 | | | | |
| Park, 2009 (100) | 21 | 30.1 | 29 | 29.8 | 90 | 15.9 | 16.7 | 33.8 | 66.4 |

*Note*: FES first-episode schizophrenia

*Table S1e: Demographic and clinical information of included studies for cortical thickness*

| Study | Control subjects | | Schizophrenia subjects | | | | | | |
|---|---|---|---|---|---|---|---|---|---|
| | n | Mean age [years] | n | Mean age [years] | Illness duration [months] | PANSS positive | PANSS negative | PANSS general | PANSS total |
| Ahmed, 2015 (101) | 31 | 39.4 | 33 | 36.4 | | | | | |
| Barry, 2019 (102) | 23 | 38.4 | 21 (non-treatment resistant) | 41.3 | 169.2 | 10.7 | 13.1 | 23.6 | 46.9 |
| | | | 21 (treatment resistant) | 41.5 | 186 | 20.5 | 19.5 | 34.9 | 76.2 |
| Besteher, 2016 (103) | 50 | 29.5 | 23 (non-suicidal) | 28.8 | | 19.9 | 19.1 | | 84.1 |
| | | | 14 (suicide attempters) | 34.4 | | 13.9 | 20.1 | | 69.2 |
| Boos, 2012 (104) | 122 | 27.5 | 155 | 26.91 | 48.24 | 15.34 | 15.41 | | 62.22 |
| Cai, 2022 (105) | 40 | 13.6 | 60 | 14.27 | 5.43 | 21.43 | 19.92 | 37.83 | 79.18 |
| Green, 2016 (106) | 22 | 39.57 | 22 | 38.86 | 165.36 | 18.29 | 17.43 | 38.19 | 73.9 |
| Gutiérrez-Galve, 2010 (107) | 38 | 25 | 37 | 26.8 | | | | | |
| Huang, 2025 (108) | 169 | 23.6 | 920 | 24.73 | 144 | 20.25 | 20.84 | 40.19 | 81.45 |
| Jamea, 2021 (106) | 15 | 28.8 | 15 | 33.9 | | | | | |
| Jessen, 2019 (107) | 59 | 24.59 | 56 | 25.04 | | 20.15 | 20.76 | 42.15 | 83.05 |
| Li, 2025 (108) | 48 | 45.04 | 44 (deficit) | 47.82 | 307.08 | 11.36 | 23.48 | 29.89 | 64.75 |
| | | | 50 (non-deficit) | 47.5 | 301.44 | 10.74 | 14.74 | 25.32 | 50.8 |
| Madre, 2020 (109) | 127 | 39 | 128 | 41 | 216 | 13 | 17 | | |
| Rosa, 2021 (110) | 243 | 30.72 | 135 (FEP) | 27.71 | 11.38 | 13.32 | 14.95 | 28.11 | 56.51 |
| | 137 | 33.24 | 240 (chronic) | 34.87 | 157.98 | 13.16 | 17.48 | 28.91 | 59.52 |
| Storvestre, 2019 (111) | 19 | 33.2 | 17 | 34.3 | | 16.3 | 14.8 | 28.9 | |
| | | | 11 (violent) | 33.2 | | 18.3 | 19.4 | 35 | |

| | | | | | | | | | |
|---|---|---|---|---|---|---|---|---|---|
| Takayanagi, 2020 (112) | 79 | 24.3 | 102 | 25.5 | 264 | | | | |
| Thormodsen, 2013 (113) | 32 | 15.9 | 22 | 16.2 | | 14.4 | 12.7 | 29.9 | |
| Voets, 2008 (114) | 25 | 16 | 25 | 16 | 16.8 | | | | |
| Wei, 2022 (115) | 98 | 26.53 | 117 | 24.73 | 15.77 | 16.16 | 18.9 | 46.13 | 91.48 |
| Xiao, 2015 (116) | 128 | 26.13 | 128 | 24.26 | 11.49 | 25.34 | 18.44 | 46.32 | 96.39 |
| Zhang, 2022 (117) | 48 | 24.81 | 44 | 24.66 | | 20.77 | 16.48 | 38.07 | 75.32 |
| Zugmann, 2013 (118) | 80 | 33.46 | 61 (treatment-resistant) | 33.8 | 154.8 | 14.1 | 18.67 | 30.39 | 63.41 |
| | | | 67 (non-treatment-resistant) | 35.81 | 146.52 | 11.61 | 15.39 | 27.73 | 54.58 |

*Note*: FEP first-episode psychosis

*Table S1e: Demographic and clinical information of included studies for gyrification*

| Study | Control subjects | | Schizophrenia subjects | | | | | | |
|---|---|---|---|---|---|---|---|---|---|
| | n | Mean age [years] | n | Mean age [years] | Illness duration [months] | PANSS positive | PANSS negative | PANSS general | PANSS total |
| Madeira, 2020 (8) | 20 | 31.5 | 20 | 31.5 | 72 | | | | |
| Madre, 2020 (109) | 127 | 39 | 128 | 41 | 216 | 13 | 17 | | |
| Palaniyappan, 2012 (10) | 41 | 28 | 57 | 26.1 | | | | | |
| Palaniyappan, 2013 (11) | 19 | 15.7 | 18 | 16.1 | 16.56 | 22.67 | 15.67 | | |
| Palaniyappan, 2014 (12) | 34 | | 39 | | | | | | |
| Plonka, 2020 (123) | 30 | 41.8 | 30 | 41.88 | 206.4 | 11.3 | 16.9 | | 61.23 |
| Rosa, 2021 (110) | 137 | 33.24 | 240 | 34.87 | 157.98 | 13.16 | 17.48 | 28.91 | 59.52 |
| Sasabayashi, 2020 (124) | 77 | 24.2 | 101 | 25.6 | 43 | | | | |
| Schultz, 2010 (125) | 54 | 26.6 | 54 | 26.4 | 2.5 | 18.3 | 18.3 | | 75.9 |
| Takayanagi, 2019 (126) | 50 | 25.8 | 37 (deficit) | 27.2 | 50.4 | | | | |
| | | | 36 (non-deficit) | 26.6 | 46.8 | | | | |
| Yunzhi, 2022 (127) | 106 | 23.67 | 108 | 24.97 | 15.13 | | | | |

*Table S1g: Demographic and clinical information of included studies for GMV*

| Study | Control subjects | | Schizophrenia subjects | | | | | | |
|---|---|---|---|---|---|---|---|---|---|
| | n | Mean age [years] | n | Mean age [years] | Illness duration [months] | PANSS positive | PANSS negative | PANSS general | PANSS total |
| Adamu, 2023 (128) | 74 | 35.82 | 72 | 38.17 | 192.36 | 14.96 | 14.53 | 29.22 | |
| Amann, 2016 (129) | 45 | 43.3 | 45 | 43.2 | 258.00 | 13.3 | 17.8 | | |
| Ananth, 2002 (130) | 20 | 38.6 | 20 | 37.8 | 190.20 | | | | |
| Anderson, 2015 (131) | 20 | 33.3 | 18 (first-line antipsychotics) | 32.2 | | | | | |
| | | | 19 (treatment-resistant) | 33.3 | 156.00 | 11 | 18 | 28 | 57 |
| | | | 15 (ultra treatment-resistant) | 34.3 | 136.80 | 13 | 20 | 29 | 62 |
| Arachchi, 2020 (132) | 116 | 33.41 | 116 | 33.75 | 116.6 | 16.8 | 20 | 34.3 | 71.1 |
| Asami, 2012 (133) | 36 | 22.9 | 33 | 22.5 | 4.88 | | | | |
| Bagary, 2003 (134) | 30 | 28.9 | 30 | 27.3 | 63.96 | | | | |
| Barsic, 2021 (135) | 50 | 35.84 | 57 (onset after age of 26) | 44.58 | 13.79 | 26.14 | 28.6 | 50.67 | 106 |
| | | | 43 (onset before age of 26) | 29.42 | 9.72 | 27.61 | 27.54 | 50.96 | 105.81 |
| Biondi, 2025 (136) | 91 | 38.54 | 74 | 37.43 | 177.84 | 14.99 | 15.15 | 28.57 | |
| Bonilha, 2008 (137) | 13 | 35 | 14 | 40 | | 24.45 | 21.27 | 43.72 | 96.63 |
| Brown, 2011 (138) | 21 | 45 | 17 | 44.8 | 229.20 | 12.3 | 13.5 | 30.2 | |
| Cai, 2022 (102) | 40 | 13.6 | 60 | 14.27 | | 21.43 | 19.92 | 37.83 | 79.18 |
| Cancel, 2015 (139) | 30 | 32.9 | 21 | 32.1 | 119.76 | | | | |
| Cascella, 2010 (140) | 90 | 46.3 | 31 (non-deficit) | 44.4 | 19.70 | | | | |
| | | | 19 (deficit) | 35.1 | 11.80 | | | | |

| | | | | | | | | | |
|---|---|---|---|---|---|---|---|---|---|
| Chen, 2014 (141) | 86 | 25.03 | 86 | 24.52 | 11.07 | 26.11 | 19.73 | 50.19 | |
| Chow, 2011(142) | 34 | 27.8 | 29 | 30.7 | 99.60 | 16.8 | 18.5 | 34.8 | |
| Cui, 2011 (143) | 36 | 26.56 | 23 | 24.78 | 48.48 | 22.48 | 20.91 | | |
| Donohoe, 2011 (144) | 19 (ZNF804A genotype AA) | 32.49 | 27 (ZNF804A genotype AA) | 40.36 | 216.24 | 25.38 | 28.38 | 60.77 | |
| | 14 (ZNF804A genotype AC) | 31.93 | 30 (ZNF804A genotype AC) | 42.71 | 209.04 | 23.63 | 22.47 | 53.3 | |
| | 5 (ZNF804A genotype CC) | 38.8 | 13 (ZNF804A genotype CC) | 42.36 | 205.08 | 24.79 | 26.18 | 55.96 | |
| Douaud, 2007 (145) | 25 | 16.008 | 25 | 16.332 | 16.80 | 22 | 16 | | |
| Egashira, 2014 (146) | 41 | 58.8 | 22 (LOS) | 58.3 | 62.40 | 8.2 | 8.4 | 18.1 | |
| | | | 24 (EOS) | 58.2 | 415.20 | 11.8 | 19 | 28.4 | |
| Euler, 2009 (147) | 23 | 43.3 | 20 | 43.26 | | | | | |
| Farrow, 2005 (148) | 13 (male) | 20 | 18 (male) | 20 | 7.00 | 18 | 20 | 39 | |
| | 9 (female) | 21 | 7 (female) | 19 | 3.00 | 18 | 18 | 44 | |
| Ferri, 2012 (149) | 19 | 28.7 | 18 | 27.2 | | 13.6 | 12 | 23.7 | |
| Frascarelli, 2015 (150) | 24 | 30.3 | 18 (<= 10 years illness duration) | 29.2 | 58.80 | 14.9 | 21.1 | 37.4 | 73.5 |
| | | | 15 (> 10 years illness duration) | 42.7 | 206.40 | 15.3 | 24.9 | 40.6 | 80.8 |
| Frascarelli, 2023 (151) | 19 | 29.7 | 16 (<= 10 years illness duration) | 43.1 | 61.2 | 15.9 | 22.2 | 39.4 | 77.5 |
| | | | 14 (> 10 years illness duration) | 29.4 | 58.8 | 15.3 | 24.4 | 40.1 | 79.8 |
| Guo, 2014a (152) | 19 | 25.16 | 19 | 23.95 | 9.37 | 19.47 | 20.26 | 38.05 | 77.79 |
| Guo, 2014b (153) | 43 | 23.67 | 20 | 23.4 | 12.85 | 22.8 | 22.2 | | 88.9 |
| Guo, 2015 (154) | 50 | 23.48 | 49 | 22.69 | 22.45 | 22.27 | 22.82 | | 91.31 |

| | | | | | | | | | |
|---|---|---|---|---|---|---|---|---|---|
| Guo, 2019(155) | 33 | 23.8 | 33 | 24.3 | | 22.4 | 20.8 | 44.5 | 88.1 |
| Henze, 2011 (156) | 13 | 17.62 | 13 | 17.09 | 7.55 | | | | |
| Herold, 2009 (157) | 21 | 27.4 | 18 | 28.7 | 40.80 | 14.2 | 19.6 | 34.6 | 68.3 |
| Horn, 2010 (158) | 20 | | 20 | 30.1 | | | | 69.85 | |
| Hu, 2013 (159) | 62 | 23.3 | 55 | 22.29 | 11.14 | 22.43 | 22.82 | | 91.49 |
| Huang, 2015 (160) | 18 | 25.06 | 18 | 22.56 | 9.19 | 24.86 | 23.92 | 48.475 | 97.25 |
| Huang, 2017 (161) | 26 | 23.15 | 18 (no delusions) | 22.5 | 8.42 | | | | |
| | | | 24 (with delusions) | 24.25 | 9.28 | | | | |
| Ivelva, 2012 (162) | 10 | 43.9 | 19 | 39.89 | | | | | |
| Jayakumar, 2005 (163) | 18 | 25.7 | 18 | 24.9 | 10.30 | 19 | 23 | 36 | 79 |
| Jiang, 2019 (164) | 20 | 41.6 | 20 | 40.3 | | 12.9 | 18 | 27.8 | 58.7 |
| John, 2015 (165) | 45 | 26.84 | 45 | 28.13 | 24.42 | 15.09 | 15.16 | 26.53 | 56.78 |
| Kasparek, 2007 (166) | 28 | 24.1 | 22 | 23.7 | 9.60 | 9.1 | 15.4 | 26.7 | 51.2 |
| Katz, 2016 (167) | 32 | 29.84 | 24 | 31.21 | | | | | 67.21 |
| Kawada, 2009 (168) | 26 | 36.3 | 26 | 36.7 | 118.80 | 14.5 | 15.5 | 31.1 | 61.1 |
| Kawasaki, 2007 (169) | 30 | 25.4 | 30 | 24.7 | 48.00 | | | | |
| Kim, 2017 (170) | 22 | 31.6 | 22 | 31.7 | 110.4 | 18.4 | 21.1 | | 39.6 |
| Lee, 2020 (171) | 65 | 34.52 | 65 | 36.98 | 186.24 | | | | |
| Lei, 2015 (172) | 44 | 22.55 | 44 (non-deficit) | 23.16 | 8.83 | 24.67 | 16.7 | 46.27 | 88.9 |
| | | | 44 (deficit) | 22.91 | 19.89 | 21.95 | 27.57 | 46.43 | 95.95 |
| Lei, 2019 (173) | 32 | 21.59 | 14 | 21.79 | 13.32 | 20.93 | 26.86 | 48.43 | 96.21 |
| Li, 2020 (174) | 86 | 24 | 86 | 23.5 | 14.2 | 21.7 | 20.6 | 44.1 | 86.8 |
| Li, 2022 (175) | 79 | 14.3 | 55 | 14.9 | | 14.9 | 13.9 | 31.7 | 65.4 |
| Liao, 2015 (176) | 99 | 25.8 | 93 | 27 | 54.10 | 23.4 | 18.3 | 35.7 | 77.3 |
| Liu, 2020 (177) | 28 | 26.07 | 48 (violent) | 28.98 | 86.04 | 25 | 17.13 | | 84.65 |
| | | | 27 (non-violent) | 27.7 | 74.76 | 21.3 | 19.89 | | 83.78 |
| Lu, 2024 (178) | 53 | 31.17 | 45 (violent) | 30.82 | 75.36 | 14.98 | 17.44 | | |

| | | | | | | | | | |
|---|---|---|---|---|---|---|---|---|---|
| | | | 45 (non-violent) | 27.93 | 57.09 | 14.73 | 14.93 | | |
| Lui, 2009 (179) | 68 | 24.7 | 68 | 24.2 | 8.60 | 26.6 | 20.6 | 51.8 | 107.4 |
| Madeira, 2020 (119) | 20 | 31.5 | 20 | 31.5 | 72 | | | | |
| McDonald, 2005 (180) | 52 | 39.3 | 25 | 37.3 | 208.80 | | | | |
| McIntosh, 2004 (181) | 49 | 35.27 | 26 | 36.85 | | | | | |
| Meda, 2008 (182) | 200 | 40.04 | 200 | 39.694 | | | | | |
| Meisenzahl, 2008 (183) | 177 | 31.5 | 93 | 28.2 | 9.10 | 19.9 | 20.4 | | 81.1 |
| Nakamura, 2013 (184) | 51 | 23.9 | 34 | 24.7 | | | | | |
| Oertel-Knöchel, 2012 (185) | 37 | 39.36 | 31 | 38 | 164.52 | 15.69 | 15.13 | 32.06 | |
| Onay, 2017 (186) | 16 | 34.4 | 20 | 36.5 | 128.4 | 38.2 | 20.1 | | |
| Ota, 2017 (187) | 62 | 40.6 | 37 | 36.2 | 162 | 15.2 | 17.2 | 32.7 | 65.1 |
| Palaniyappan, 2012 (188) | 41 | 28.04 | 57 | 26.1 | | | | | |
| Picado, 2015 (189) | 20 | 33.2 | 20 | 35.9 | | 12.25 | 15.5 | | 59.35 |
| Poeppl, 2014 (190) | 30 | 30.2 | 43 | 27.75 | | 21.65 | 26.5 | 49.95 | 95.1 |
| Poletti, 2016 (191) | 136 | 33.31 | 96 | 37.54 | 151.32 | 17.62 | 20.41 | 12.61 | |
| Pomarol-Clotet, 2012 (192) | 32 | 41.03 | 32 | 41.56 | 261.48 | | | | 71.97 |
| Price, 2010 (193) | 47 | 24.8 | 48 | 26.2 | | | | | |
| Rametti, 2012 (194) | 23 | 31.6 | 23 | 32.1 | 126.00 | 17.2 | 22.3 | 37.7 | 77.3 |
| Rose, 2014 (195) | 150 | 33.49 | 163 | 38.51 | 180.00 | | | | |
| Salgado-Pineda, 2004 (196) | 12 | 25.14 | 14 | 25.05 | | | | | |
| Salgado-Pineda, 2011 (197) | 14 | 34.6 | 14 | 37.3 | 168.00 | 24.7 | 13.1 | | 60.7 |

| | | | | | | | | | |
|---|---|---|---|---|---|---|---|---|---|
| Salgado-Pineda, 2014 (198) | 14 | 34.64 | 14 | 37.29 | 168.00 | 24.71 | 13.14 | | 60.7 |
| Sanjuán, 2020 (199) | 18 | 38.24 | 61 | 31.87 | | 15.8 | 16.5 | 31.98 | 64.24 |
| Sans-Sansa, 2013 (200) | 59 | 38.31 | 20 | 46.1 | 286.08 | | | | |
| Sarró, 2013 (201) | 61 | 40.69 | 32 (tardive dyskinesia) | 43.09 | 268.32 | 14.45 | 17.1 | | 76.07 |
| | | | 49 (non-tardive dyskinesia) | 42.82 | 249.72 | 16 | 19.94 | | 73.57 |
| Schiffer, 2010 (202) | 14 | 36.7 | 12 (with alcohol misuse) | 37.8 | 201.60 | 14.6 | 20.5 | 31.9 | |
| | | | 12 (without alcohol misuse) | 37.2 | 139.20 | 16 | 18.7 | 35.8 | |
| Schuster, 2012 (203) | 40 | 62.2 | 27 | 59.9 | 350.40 | 20 | 25.7 | | 64 |
| Shen, 2022 (204) | 25 | 12.68 | 20 | 13.95 | | 17.07 | 21.07 | 38.93 | 77.07 |
| Singh, 2018 (205) | 28 | 31.44 | 28 | 33.89 | 479.65 | | | | |
| Spalthoff, 2018 (206) | 102 | 33.15 | 51 | 35.18 | 105.6 | | | | |
| Spaniel, 2011 (207) | 36 | 25.1 | 40 | 32.3 | 40.80 | 10.3 | 12.5 | 25 | 47.8 |
| Stegmayer, 2014 (208) | 34 | 37.1 | 43 | 34.1 | 104.40 | 15.4 | 14.6 | | 57.4 |
| Sungur, 2025 (209) | 86 | 37.41 | 75 (subtype 1) | 37.4 | 170.04 | 11.36 | 19.78 | 30.21 | 61.21 |
| | | | 61 (subtype 2) | 37.6 | 179.64 | 10.92 | 20.63 | 29.39 | 60.92 |
| Szendi, 2017 (210) | 13 | 34 | 21 | 39 | | | | | |
| Tan, 2015 (211) | 17 | 41.2 | 18 | 40.5 | 190.80 | 8.7 | 21.7 | | 59.1 |
| Tang, 2012 (212) | 34 | 16.6 | 29 | 16.5 | 9.30 | 22.3 | 20.8 | 34.5 | 77.7 |
| Tomelleri, 2009 (213) | 79 | 40.29 | 70 | 39.73 | 169.56 | | | | |
| Torres, 2016 (214) | 151 | 30.6 | 161 | 30.4 | 60.00 | | | | 56.1 |
| Tregellas, 2007 (215) | 32 | 35.3 | 32 | 39.6 | | | | | |

| | | | | | | | | | |
|---|---|---|---|---|---|---|---|---|---|
| Tseng, 2021 (216) | 26 | 35.62 | 37 | 35.7 | 146.28 | 13.43 | 16.65 | 25.68 | 55.76 |
| van Tol, 2014 (217) | 51 | 36.14 | 51 | 34.04 | 105.24 | 11.48 | 14.37 | 29.84 | |
| Venkatasubramanian, 2010 (218) | 27 | 27.4 | 30 | 30.1 | 41.70 | 22 | 23 | 39 | |
| Watson, 2012 (219) | 25 | 28.2 | 25 | 28.8 | | | | | |
| Whitford, 2006 (220) | 47 | 19.3 | 41 | 19.8 | 8.30 | 17.7 | 18.8 | 40.3 | |
| Witthaus, 2009 (221) | 29 | 25.7 | 23 | 26.4 | | 19.3 | 18 | | |
| Wu, 2022 (222) | 54 (OWB) | 33.31 | 69 (OWB) | 33.88 | 93.72 | 21.38 | 21.25 | 39 | 81.62 |
| | 53 (NW) | 31.42 | 74 (NW) | 31.86 | 85.2 | 21.53 | 20.26 | 36.66 | 78.45 |
| Xie, 2021 (54) | 33 | 32.03 | 30 | 30.3 | 21.36 | 21.36 | 19.65 | 19.85 | 40.35 |
| Yamada, 2007 (223) | 20 | 39.1 | 20 | 38.8 | 139.2 | 16.4 | 15.7 | 32.4 | 64.5 |
| Yang, 2022 (224) | 95 | 30.21 | 70 | 28.4 | 43.26 | 22.74 | 19.86 | | 78.5 |
| Yokoyama, 2018 (225) | 40 | 35.05 | 60 | 38.03 | | 13.785 | 15.865 | 30.2 | |
| Yue, 2016 (226) | 24 | 24.79 | 20 | 24.45 | 22.92 | 21.25 | 19.1 | 48.6 | 88.95 |
| Yüksel, 2012 (227) | 45 | 36.4 | 61 | 38.7 | | 18.1 | | | 62.3 |
| Zhang, 2015 (228) | 30 | 15.3 | 37 | 15.5 | 16.00 | 20.42 | 20.91 | 33.28 | 74.62 |
| Zhang, 2017 (229) | 83 | 27.84 | 75 | 23.71 | 139.68 | | | | |
| Zierhut, 2013 (230) | 36 | 30.97 | 34 | 34.59 | 120.36 | 16.39 | 28.97 | 35.73 | 71.09 |

*Note*: EOS early-onset schizophrenia, LOS late-onset schizophrenia, NW normal weight, OWB overweight/obesity

**Supplementary Tables S2: Peak coordinates of single-modality meta-analyses at a significance level of p<0.001**

*Table S2a: Peak coordinates of ALFF meta-analysis*

| MNI coordinates | SDM-Z | p-value | voxels | description |
|---|---|---|---|---|
| **Positive** | | | | |
| 24,6,-12 | 5.504 | ~0 | 463 | (undefined), BA 48 |
| -16,14,8 | 4.738 | 1.073E-06 | 448 | Left anterior thalamic projections |
| 44,42,-12 | 5.23 | 0.00000006 | 405 | Right inferior frontal gyrus, orbital part, BA 47 |
| -38,40,-6 | 3.769 | 8.2016E-05 | 106 | Left anterior thalamic projections |
| 18,12,10 | 3.496 | 0.00023609 | 53 | Right anterior thalamic projections |
| 48,22,12 | 4.357 | 6.557E-06 | 46 | Right inferior frontal gyrus, triangular part, BA 45 |
| **Negative** | | | | |
| -54,-16,38 | -6.202 | ~0 | 650 | Left postcentral gyrus, BA 3 |
| 52,-22,44 | -5.949 | ~0 | 682 | Right postcentral gyrus, BA 3 |
| -20,-68,-14 | -4.359 | 6.557E-06 | 124 | Left cerebellum, hemispheric lobule VI, BA 18 |
| -6,50,-16 | -4.005 | 3.0994E-05 | 115 | Corpus callosum |
| -4,-40,64 | -4.163 | 1.5676E-05 | 48 | Left precuneus, BA 5 |
| 2,-60,16 | -4.013 | 2.9922E-05 | 34 | Right calcarine fissure / surrounding cortex |
| 18,-84,26 | -3.677 | 0.0001182 | 19 | Corpus callosum |

*Table S2b: Peak coordinates of ReHo meta-analysis*

| MNI coordinates | SDM-Z | p-value | voxels | description |
|---|---|---|---|---|
| **Positive** | | | | |
| 20,6,-2 | 5.288 | 0.00000006 | 413 | Right striatum |
| -18,6,-6 | 5.281 | 0.00000006 | 153 | Left striatum |
| 14,-2,20 | 4.604 | 2.086E-06 | 82 | Right caudate nucleus |
| 0,42,38 | 4.275 | 9.537E-06 | 68 | Left superior frontal gyrus, medial, BA 9 |
| -40,16,6 | 3.864 | 5.5671E-05 | 53 | Left insula, BA 48 |
| -16,8,14 | 3.715 | 0.00010163 | 29 | Left anterior thalamic projections |
| 46,46,-6 | 3.762 | 8.4102E-05 | 25 | Right inferior frontal gyrus, orbital part, BA 46 |
| -44,-22,-30 | 3.466 | 0.00026387 | 17 | Left inferior temporal gyrus, BA 20 |
| 4,32,54 | 3.626 | 0.00014412 | 16 | Right superior frontal gyrus, medial |
| **Negative** | | | | |
| 0,48,-10 | -7.121 | ~0 | 1226 | Left superior frontal gyrus, medial orbital, BA 11 |
| -60,-16,36 | -6.188 | ~0 | 953 | Left postcentral gyrus, BA 43 |
| -48,-76,2 | -5.996 | ~0 | 584 | Left middle occipital gyrus, BA 19 |
| 4,-42,58 | -5.882 | ~0 | 445 | Right precuneus, BA 5 |
| 54,-12,36 | -4.789 | 8.34E-07 | 397 | Right postcentral gyrus, BA 3 |
| 50,-62,-10 | -5.171 | 1.19E-07 | 295 | Right inferior temporal gyrus, BA 37 |
| 12,-76,44 | -3.856 | 5.7757E-05 | 36 | Right precuneus, BA 7 |
| 54,-32,44 | -3.362 | 0.00038743 | 11 | Right supramarginal gyrus, BA 2 |

*Table S2c: Peak coordinates of rCBF meta-analysis*

| MNI coordinates | SDM-Z | p-value | voxels | description |
|---|---|---|---|---|
| **Positive** | | | | |
| -28,4,10 | 4.589 | 2.205E-06 | 117 | Left lenticular nucleus, putamen, BA 48 |
| **Negative** | | | | |
| -46,46,-4 | -5.218 | 1.19E-07 | 481 | Left inferior frontal gyrus, orbital part, BA 46 |
| -2,46,12 | -3.945 | 3.9816E-05 | 62 | Left anterior cingulate / paracingulate gyri, BA 32 |
| -46,12,36 | -3.786 | 7.6592E-05 | 47 | Left middle frontal gyrus, BA 44 |
| -6,42,28 | -3.912 | 4.5836E-05 | 44 | Left superior frontal gyrus, medial, BA 32 |
| 36,-90,6 | -3.506 | 0.00022709 | 35 | Right middle occipital gyrus, BA 18 |

*Table S2d: Peak coordinates of glucose metabolism meta-analysis*

| MNI coordinates | SDM-Z | p-value | voxels | description |
|---|---|---|---|---|
| **Positive** | | | | |
| 48,-2,-40 | 3.691 | 0.00011164 | 18 | Right inferior temporal gyrus, BA 20 |
| **Negative** | | | | |
| 0,36,36 | -3.791 | 0.000075042 | 135 | Left superior frontal gyrus, medial, BA 9 |

*Table S2e: Peak coordinates of CTh meta-analysis*

| MNI coordinates | SDM-Z | p-value | voxels | description |
|---|---|---|---|---|
| **Negative** | | | | |
| 40,-18,0 | -3.658 | 0.00012702 | 24 | Right insula, BA 48 |

*Table S2f: Peak coordinates of gyrification meta-analysis*

None.

*Table S2g: Peak coordinates of GMV meta-analysis*

| MNI coordinates | SDM-Z | p-value | voxels | description |
|---|---|---|---|---|
| **Negative** | | | | |
| -50,-8,4 | -12.056 | ~0 | 32832 | Left superior temporal gyrus |
| 52,12,-12 | -11.034 | ~0 | 16422 | Right temporal pole, superior temporal gyrus, BA 38 |
| -4,-8,10 | -6.683 | ~0 | 1147 | Left thalamus |
| 40,-50,-22 | -6.225 | ~0 | 1333 | Right fusiform gyrus, BA 37 |
| -34,56,2 | -5.488 | ~0 | 403 | Left middle frontal gyrus, BA 10 |
| 50,-66,30 | -6.003 | ~0 | 301 | Right angular gyrus, BA 39 |
| -18,-34,0 | -3.928 | 4.2856E-05 | 14 | Left hippocampus, BA 27 |

**Supplementary Tables S3: Heterogeneity and potential publication bias**

*Table S3a: Heterogeneity and potential publication bias for ALFF*

| MNI coordinates | $I^2$-Statistic | Egger's test p-value |
|---|---|---|
| **Positive** | | |
| 24,6,-12 | 2.92 | 0.979 |
| -16,14,8 | 0.33 | 0.371 |
| 44,42,-12' | 2.64 | 1 |
| -38,40,-6' | 9.88 | 1 |
| 18,12,10 | **71.20** | 0.878 |
| 48,22,12 | 6.31 | 0.992 |
| **Negative** | | |
| -54,-16,38 | 1.56 | 0.938 |
| 52,-22,44 | 10.39 | 0.992 |
| -20,-68,-14 | 1.77 | 0.967 |
| -6,50,-16' | 10.34 | 0.967 |
| -4,-40,64 | 2.76 | 0.999 |
| 2,-60,16 | 2.07 | 0.982 |
| 18,-84,26 | 24.35 | 0.869 |

*Table S3b: Heterogeneity and potential publication bias for ReHo*

| MNI coordinates | $I^2$-Statistic | Egger's test p-value |
|---|---|---|
| **Positive** | | |
| 20,6,-2 | 24.29 | 0.426 |
| -18,6,-6 | 19.86 | 1 |
| 14,-2,20 | 2.44 | 0.989 |
| 0,42,38 | 6.37 | 0.595 |
| -40,16,6 | 3.4 | 0.739 |
| -16,8,14 | 21.32 | 0.882 |
| 46,46,-6 | 14.61 | 0.828 |
| -44,-22,-30 | 10 | 0.892 |
| 4,32,54 | 7.51 | 1 |
| **Negative** | | |
| 0,48,-10 | 0.2 | 0.918 |
| -60,-16,36 | 5 | 0.998 |
| -48,-76,2 | 23.15 | 0.829 |
| 4,-42,58 | 9.73 | 0.916 |
| 54,-12,36 | 9.59 | 1 |
| 50,-62,-10 | 8.9 | 0.859 |
| 12,-76,44 | 8.59 | 0.782 |
| 54,-32,44 | 29.62 | 0.634 |

*Table S3c: Heterogeneity and potential publication bias for rCBF*

| MNI coordinates | $I^2$-Statistic | Egger's test p-value |
|---|---|---|
| **Positive** | | |
| -28,4,10 | 2.35 | 0.908 |
| **Negative** | | |
| -46,46,-4 | 6.01 | 0.959 |
| -2,46,12 | 1.29 | 0.999 |
| -46,12,36 | 6.89 | 0.294 |
| -6,42,28 | 1.87 | 0.949 |
| 36,-90,6 | 20.58 | **<0.001** |

*Table S3d: Heterogeneity and potential publication bias for glucose metabolism*

| MNI coordinate | $I^2$-Statistic | Egger's test p-value |
|---|---|---|
| **Positive** | | |
| 48,-2,-40 | 4.76 | n.a. |
| **Negative** | | |
| 0,36,36 | 3.98 | n.a. |

Note: Egger’s test could not be conducted due to small number of studies.

*Table S3e: Heterogeneity and potential publication bias for CTh*

| MNI coordinate | $I^2$-Statistic | Egger's test p-value |
|---|---|---|
| **Negative** | | |
| 40,-18,0 | 3.97 | 0.655 |

*Table S3f: Heterogeneity and potential publication bias for gyrification*

None.

*Table S3g: Heterogeneity and potential publication bias for GMV*

| MNI coordinate | $I^2$-Statistic | Egger's test p-value |
|---|---|---|
| **Negative** | | |
| -50,-8,4 | 3.11 | 1 |
| 52,12,-12 | 4.22 | 1 |
| -4,-8,10 | 17.05 | 1 |
| 40,-50,-22 | 1.65 | **<0.001** |
| -34,56,2 | 24.87 | 1 |
| 50,-66,30 | 9.78 | 0.668 |
| -18,-34,0 | 14.45 | 0.953 |

**Supplementary Table S4: Clusters of multimodal brain abnormalities in schizophrenia for K=2 component=2 with threshold 0.0075**

| Cluster index | $n_{voxels}$ | Maximum value | MNI peak coordinates | | | Harvard-Oxford regions |
|---|---|---|---|---|---|---|
| | | | x | y | z | |
| 19 | 10358 | 0.00929 | 18 | 18 | -12 | Frontal pole (14.4%), insular cortex (13.2%), central opercular cortex (9.7%) |
| 18 | 8120 | 0.00961 | -26 | 6 | 8 | Insular cortex (17.0%), temporal pole (12.9%), central opercular cortex (9.4%) |
| 17 | 1483 | 0.00847 | 6 | 36 | 54 | Cingulate gyrus (anterior division, 39.7%), superior frontal gyrus (38.0%), paracingulate gyrus (20.5%) |
| 16 | 104 | 0.0081 | 6 | -36 | 26 | Cingulate gyrus (posterior division, 100.0%) |
| 15 | 22 | 0.00781 | -8 | -22 | 38 | Cingulate gyrus (posterior division, 100.0%) |
| 14 | 18 | 0.00789 | -44 | 32 | 2 | Frontal pole (66.7%), inferior frontal gyrus (pars triangularis, 33.3%) |
| 13 | 18 | 0.00785 | -26 | 14 | 44 | Middle frontal gyrus (100.0%) |
| 12 | 17 | 0.00807 | -20 | 32 | -18 | Frontal orbital cortex (88.2%), frontal pole (11.8%) |
| 11 | 15 | 0.00786 | 70 | -28 | -12 | Middle temporal gyrus (posterior division, 100.0%) |
| 10 | 10 | 0.00781 | 34 | 36 | 36 | Frontal pole (90.0%), middle frontal gyrus (10.0%) |
| 9 | 10 | 0.0078 | -48 | 26 | 0 | Frontal operculum cortex (44.4%), frontal orbital cortex (33.3%), inferior frontal gyrus (pars triangularis, 22.2%) |
| 8 | 8 | 0.00775 | -44 | 38 | -4 | Frontal pole (100%) |
| 7 | 7 | 0.00783 | -36 | -30 | -18 | Temporal fusiform cortex (posterior division, 100.0%) |
| 6 | 6 | 0.0079 | -44 | 36 | 8 | Inferior frontal gyrus (pars triangularis, 100.0%) |
| 5 | 2 | 0.00788 | -36 | 38 | -6 | n.a. |
| 4 | 2 | 0.00759 | -30 | -34 | -24 | Temporal fusiform cortex (posterior division, 100.0%) |
| 3 | 2 | 0.00783 | 36 | 0 | -34 | n.a. |
| 2 | 2 | 0.00755 | -2 | 6 | 42 | Cingulate gyrus (anterior division, 100.0%) |

| | | | | | | |
|---|---|---|---|---|---|---|
| 1 | 1 | 0.00773 | -36 | -42 | -24 | Fusiform cortex (posterior division, 100.0%) |

*Note*: the top three Harvard-Oxford regions are stated.

**Supplementary Table S5: Percentage of overlap between multimodal abnormality clusters and large-scale brain networks from Yeo and colleagues**

| Cluster index | Yeo-7 networks | | | | | | |
|---|---|---|---|---|---|---|---|
| | Frontoparietal control | Default Mode | Dorsal Attention | Limbic | Salience | Somato-motor | Visual |
| 19 | 7.511 | 11.981 | 0.56 | 2.8 | 12.3 | 12.425 | 0 |
| 18 | 1.835 | 14.815 | 0.369 | 9.335 | 13.276 | 7.833 | 0 |
| 17 | 17.532 | 14.093 | 0 | 0 | 17.937 | 0 | 0 |
| 16 | 14.423 | 46.154 | 0 | 0 | 0 | 0 | 0 |
| 15 | 0 | 0 | 0 | 0 | 54.545 | 0 | 0 |
| 14 | 0 | 0 | 0 | 0 | 0 | 0 | 0 |
| 13 | 0 | 61.111 | 0 | 0 | 0 | 0 | 0 |
| 12 | 0 | 0 | 0 | 70.588 | 0 | 0 | 0 |
| 11 | 0 | 100 | 0 | 0 | 0 | 0 | 0 |
| 10 | 10 | 0 | 0 | 0 | 0 | 0 | 0 |
| 9 | 0 | 20 | 0 | 0 | 0 | 0 | 0 |
| 8 | 12.5 | 62.5 | 0 | 0 | 0 | 0 | 0 |
| 7 | 0 | 0 | 14.286 | 0 | 0 | 0 | 14.286 |
| 6 | 100 | 0 | 0 | 0 | 0 | 0 | 0 |
| 5 | 0 | 0 | 0 | 0 | 0 | 0 | 0 |
| 4 | 0 | 0 | 0 | 0 | 0 | 0 | 100 |
| 3 | 0 | 0 | 0 | 0 | 0 | 0 | 0 |
| 2 | 0 | 0 | 0 | 0 | 0 | 0 | 0 |
| 1 | 0 | 0 | 0 | 0 | 0 | 0 | 100 |

*Note:* Percentages do not necessarily add up to 100% since part of the clusters were outside the defined network cortical masks.

**Supplementary Table S6: Spatial correlation analysis between multi-modal macroscopic brain abnormalities and cell-type specific gene expression**

| Cell type | Spearman's ρ | $p_{spin}$ |
|---|---|---|
| Astrocytes (Astro) | **0.471** | **0.023** |
| Endothelial cells (Endo) | 0.162 | 0.369 |
| Microglia (Micro) | **0.430** | **0.035** |
| Excitatory neurons (Neuro-Ex) | 0.205 | 0.205 |
| Inhibitory neurons (Neuro-In) | 0.170 | 0.245 |
| Oligodendrocyte precursor cells (OPCs) | -0.105 | 0.500 |
| Oligodendrocytes (Oligo) | **0.549** | **0.006** |
| Pericytes (Per) | 0.075 | 0.646 |
